# Rare variation in noncoding regions with evolutionary signatures contributes to autism spectrum disorder risk

**DOI:** 10.1101/2023.09.19.23295780

**Authors:** Taehwan Shin, Janet H.T. Song, Michael Kosicki, Connor Kenny, Samantha G. Beck, Lily Kelley, Xuyu Qian, Julieta Bonacina, Frances Papandile, Irene Antony, Dilenny Gonzalez, Julia Scotellaro, Evan M. Bushinsky, Rebecca E. Andersen, Eduardo Maury, Len A. Pennacchio, Ryan N. Doan, Christopher A. Walsh

## Abstract

Little is known about the role of noncoding regions in the etiology of autism spectrum disorder (ASD). We examined three classes of noncoding regions: Human Accelerated Regions (HARs), which show signatures of positive selection in humans; experimentally validated neural Vista Enhancers (VEs); and conserved regions predicted to act as neural enhancers (CNEs). Targeted and whole genome analysis of >16,600 samples and >4900 ASD probands revealed that likely recessive, rare, inherited variants in HARs, VEs, and CNEs substantially contribute to ASD risk in probands whose parents share ancestry, which enriches for recessive contributions, but modestly, if at all, in simplex family structures. We identified multiple patient variants in HARs near *IL1RAPL1* and in a VE near *SIM1* and showed that they change enhancer activity. Our results implicate both human-evolved and evolutionarily conserved noncoding regions in ASD risk and suggest potential mechanisms of how changes in regulatory regions can modulate social behavior.

## Introduction

Autism spectrum disorder (ASD) is a highly heritable, phenotypically complex condition that affects 2-3% of children (Maenner et al., 2023) and shares comorbidity with many conditions including intellectual disability, attention-deficit/hyperactivity disorder, and obesity (Hyman et al., 2020). Over the last decade, immense progress has been made in understanding the genetic underpinnings of ASD. This has been largely driven by investigating *de novo* coding variants (De Rubeis et al., 2014; Iossifov et al., 2014; Zhou et al., 2022), and more recently, rare, recessive, inherited coding variants (Doan et al., 2019; Ruzzo et al., 2019; Zhou et al., 2022) of moderate to large effect size. Together, these efforts have identified more than 1000 candidate genes (Abrahams et al., 2013), with many identified ASD genes converging on similar gene programs, including synapse formation and maintenance, chromatin remodeling, and cytoskeletal pathways (De Rubeis et al., 2014; Ruzzo et al., 2019; Satterstrom et al., 2020).

Despite advances in understanding the role of coding variation in ASD, little is known about the role of noncoding variation. One major obstacle is that 98.5% of the genome is noncoding, and a systematic analysis of the entire noncoding genome requires a commensurately larger sample size to reach statistical significance. To address this issue, a number of studies have reduced the noncoding sequence search space to focus on noncoding regions that are likely to be functional and then queried whether specific classes of noncoding regions are enriched for patient variants. Evolutionary conservation has emerged as a strong marker of likely functional regions; many conserved noncoding regions are known to function as developmental enhancers, and disease-associated variants in these regions have been shown to disrupt gene regulation during development (Polychronopoulos et al., 2017). Indeed, recent studies found that *de novo* variants in conserved promoters are enriched in patients with ASD (An et al., 2018) and that *de novo* variants in conserved fetal brain enhancers are enriched in patients with severe neurodevelopmental disorders (Short et al., 2018). Consanguineous families, which are enriched for recessive contributions because of shared ancestry, have also proved powerful for identifying the contribution to ASD of noncoding regions, including inherited, homozygous deletions, which have not been detectable in nonconsanguineous families (Morrow et al., 2008; Schmitz-Abe et al., 2020).

Concurrently, multiple studies suggest that noncoding regions that show evolutionary signatures of selection in humans may be preferentially vulnerable in human diseases (Oksenberg et al., 2013; Xu et al., 2015; Doan et al., 2016; Srinivasan et al., 2016; Song et al., 2018). For instance, human accelerated regions (HARs) are regions that are highly conserved across species, but show signals of positive selection in the human evolutionary lineage (Pollard et al., 2006; Bird et al., 2007; Prabhakar et al., 2008; Lindblad-Toh et al., 2011; Gittelman et al., 2015; Girskis et al., 2021). HARs have been found to be enriched near genes associated with brain development (Capra et al., 2013; Kamm et al., 2013; Oksenberg et al., 2013; Boyd et al., 2015), and rare, recessive variants in HARs are enriched in patients with ASD in consanguineous families (Doan et al., 2016).

Here, we characterize three classes of noncoding regions that vary in evolutionary conservation and selection, examine their functional activity, and find that rare, inherited, likely recessive variants in all three classes are enriched, to varying degrees, in ASD cases compared to controls. We find that family structure strongly affects the contribution of rare, recessive variants across consanguineous, non-consanguineous multiplex, and non-consanguineous simplex cohorts. These ASD cohorts nominate a set of rare, recessive patient variants for further study, and we demonstrate that patient variants near the ASD-associated gene *IL1RAPL1* and the neurobehavioral gene *SIM1* regulate gene expression, suggesting that they may affect ASD risk.

## Results

### HARs, CNEs, and VEs may act as regulatory elements in the brain

Based on prior studies that suggest that regions that are highly conserved or under selection in humans may be selectively vulnerable in neurodevelopmental diseases (Doan et al., 2016; An et al., 2018; Short et al., 2018), we identified three classes of noncoding regions for characterization (Fig. 1, Table S1): (1) HARs, which are regions conserved through other mammals that are likely under positive selection in humans (Girskis et al., 2021) and which were previously shown to have elevated rates of rare, recessive variants in a consanguineous ASD cohort (Doan et al., 2016); (2) neural Vista Enhancers (VEs), which are conserved elements that have been experimentally tested to drive reporter activity in the brain in E11.5 transient transgenic reporter mice (Visel et al., 2007); and (3) conserved neural enhancers (CNEs). We defined CNEs as elements that are highly conserved across species, are highly constrained within humans, and are predicted to be enhancers in fetal brain and neurospheres or adult brain by ChromHMM from the Roadmap Epigenomics Project (Kundaje et al., 2015) (Materials and Methods).

**Figure 1.**
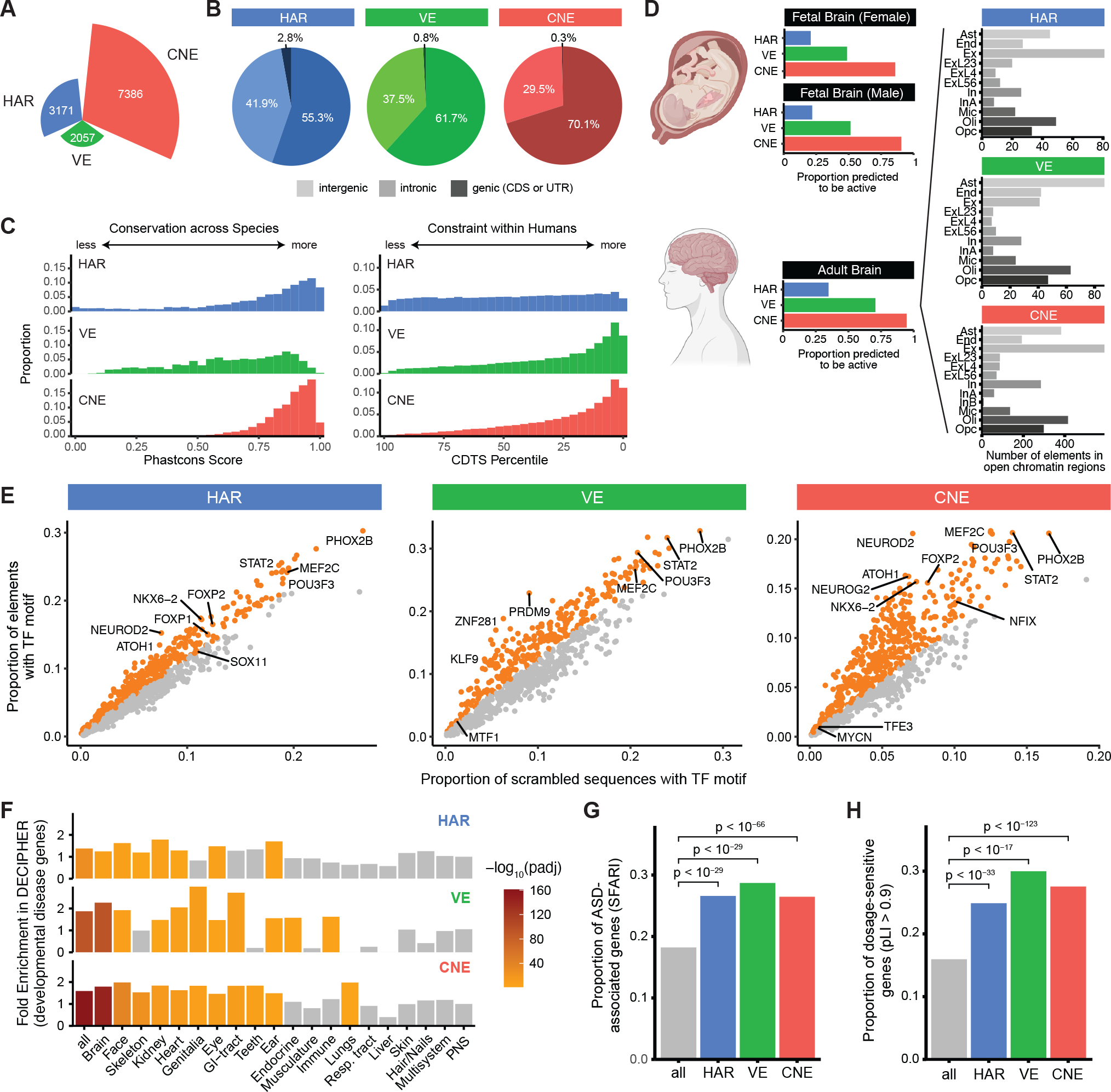
Genomic and epigenomic features of HARs, VEs, and CNEs. (A) Number of HARs, VEs, and CNEs. (B) Proportion of HARs, VEs, and CNEs in intergenic (light coloring), intronic (moderate coloring), and genic (dark coloring) regions. (C) Conservation across species (left) and constraint within humans (right) of HARs, VEs, and CNEs are represented by phastCons score (Siepel et al., 2005) and CDTS percentile (di Iulio et al., 2018), respectively. (D) Proportion of HARs, VEs, and CNEs predicted to be active by ChromHMM based on epigenomic data from a fetal male brain, a fetal female brain, and an adult brain (Kundaje et al., 2015) (left). Number of HARs, VEs, and CNEs that overlap open chromatin regions from scTHS-seq across cell types in the adult brain (Lake et al., 2018). Ast: astrocytes, End: endothelial cells, Ex: excitatory neurons, ExL23: layers 2-3 excitatory neurons, ExL4: layers 4 excitatory neurons, ExL56: layers 5-6 excitatory neurons, In: inhibitory neurons, InA: inhibitory neurons subtype A, InB: inhibitory neurons subtype B, Mic: microglia, Oli: oligodendrocytes, Opc: oligodendrocyte precursor cells. (E) Enrichment of transcription factor (TF) binding site motifs in HARs, VEs, and CNEs compared to nucleotide-matched scrambled sequences (Materials and Methods). Orange dots indicate significantly enriched elements at 5% FDR. (F) Enrichment of HARs, VEs, and CNEs near genes associated with developmental diseases in different body systems from the DECIPHER consortium (Firth et al., 2009). (G) HARs, VEs, and CNEs are enriched near ASD-associated genes annotated in the SFARI database (Abrahams et al., 2013). (H) Genes near HARs, VEs, or CNEs are enriched for genes with pLI > 0.9. Genes with pLI > 0.9 are considered loss-of-function intolerant (Lek et al., 2016).

Comparison of genomic and epigenomic features of HARs, VEs, and CNEs reveals similarities and differences in conservation, mutational constraint, and predicted functional activity. Most HARs and CNEs are highly conserved across species, whereas VEs exhibit variability in their level of conservation (Fig. 1C), likely because VEs often contain conserved segments flanked by stretches of less conserved sequences (Visel et al., 2007). In contrast, most VEs and CNEs are highly constrained within humans, whereas HARs, which could have either gained or lost functional activity in humans, exhibit variability in their levels of mutational constraint (Fig. 1C). As expected given our definition of CNEs and VEs, nearly all CNEs and the majority of VEs are predicted to be active in fetal or adult hu-man brain by ChromHMM (Kundaje et al., 2015) (Fig. 1D). However, HARs, which were defined solely from genomic sequence changes, are less likely to be predicted to be active in fetal (∼ 20%) or adult (∼35%) brain. Substantial proportions of HARs, VEs, and CNEs are also predicted to be active in other tissues by ChromHMM (Fig. S1), and have differing cell type specificity in the adult brain (Lake et al., 2018) (Fig. 1D). HARs, VEs, and CNEs are all enriched for transcription factor (TF) binding sites of known neurodevelopmental TFs (Fig. 1E), including FOXP2 in HARs and CNEs (den Hoed et al., 2021) and ZNF281 in VEs (Pieraccioli et al., 2018), although only CNEs are enriched in aggregate for the motifs of TFs involved in neural functions (Table S1).

### HARs, CNEs, and VEs are enriched near ASD-associated and dosage-sensitive genes

We might expect that if HARs, VEs, and CNEs modulate ASD risk, they would directly regulate the expression of genes previously implicated in ASD or other neurodevelopmental disorders. We find that HARs, VEs, and CNEs are enriched near genes implicated in severe developmental disorders that affect the brain, as annotated by the DECIPHER consortium (Firth et al., 2009) (Fig. 1F). We also observe a strong enrichment of HARs, VEs, and CNEs near ASD-associated genes, as annotated by SFARI (Abrahams et al., 2013) (adjusted *p* < 10^−29^; Fig. 1G).

Given the restricted effect of a single regulatory element on gene expression (Cannavò et al., 2016; Osterwalder et al., 2018), noncoding regions that contribute to ASD risk might preferentially regulate genes that are dosage-sensitive, i.e., genes where a small change in expression can lead to a phenotypic outcome. As a measure of dosage sensitivity, we examined loss-of-function intolerance (Lek et al., 2016; Karczewski et al., 2020). ASD-associated genes (SFARI), which have been primarily identified from *de novo* heterozygous coding variants (Iossifov et al., 2014; Zhou et al., 2022), are strongly enriched for dosage-sensitive genes as expected (Fig. S3). Strikingly, HARs, VEs, and CNEs are all also significantly enriched near dosage-sensitive genes (adjusted *p* < 10^−17^; Fig. 1H).

### VEs and CNEs are more likely than HARs to act as enhancers in neural cells

To directly test whether HARs, VEs, and CNEs can act as enhancers, we used a **ca**pture-based **M**assively **P**arallel **R**eporter **A**ssay (caMPRA) (Fig. 2A) (Girskis et al., 2021). Unlike conventional, oligonucleotide synthesis-based MPRA methods that can only test ∼150-200bp sequences cost-effectively, caMPRA can test thousands of long (∼500bp) noncoding sequences in parallel. This is critical because 60.6% of HARs, 34.7% of conserved cores of VEs, and 39.3% of CNEs are >200bp in length; conversely, 91.2% of HARs, 74.6% of conserved cores of VEs, and 92.9% of CNEs are <500bp in length (Materials and Methods; Fig. S4). Using this method, we tested HARs, VEs, and CNEs for regulatory activity in Neuro2A (N2A) cells, a neuroblastoma cell line that has been previously used to assess the neural function of noncoding regions (Doan et al., 2016; Mulvey and Dougherty, 2021; Arora et al., 2022; Mikl et al., 2022) (Materials and Methods). Enhancer activity was highly correlated across replicates (Fig. S5, Fig. S6F).

**Figure 2.**
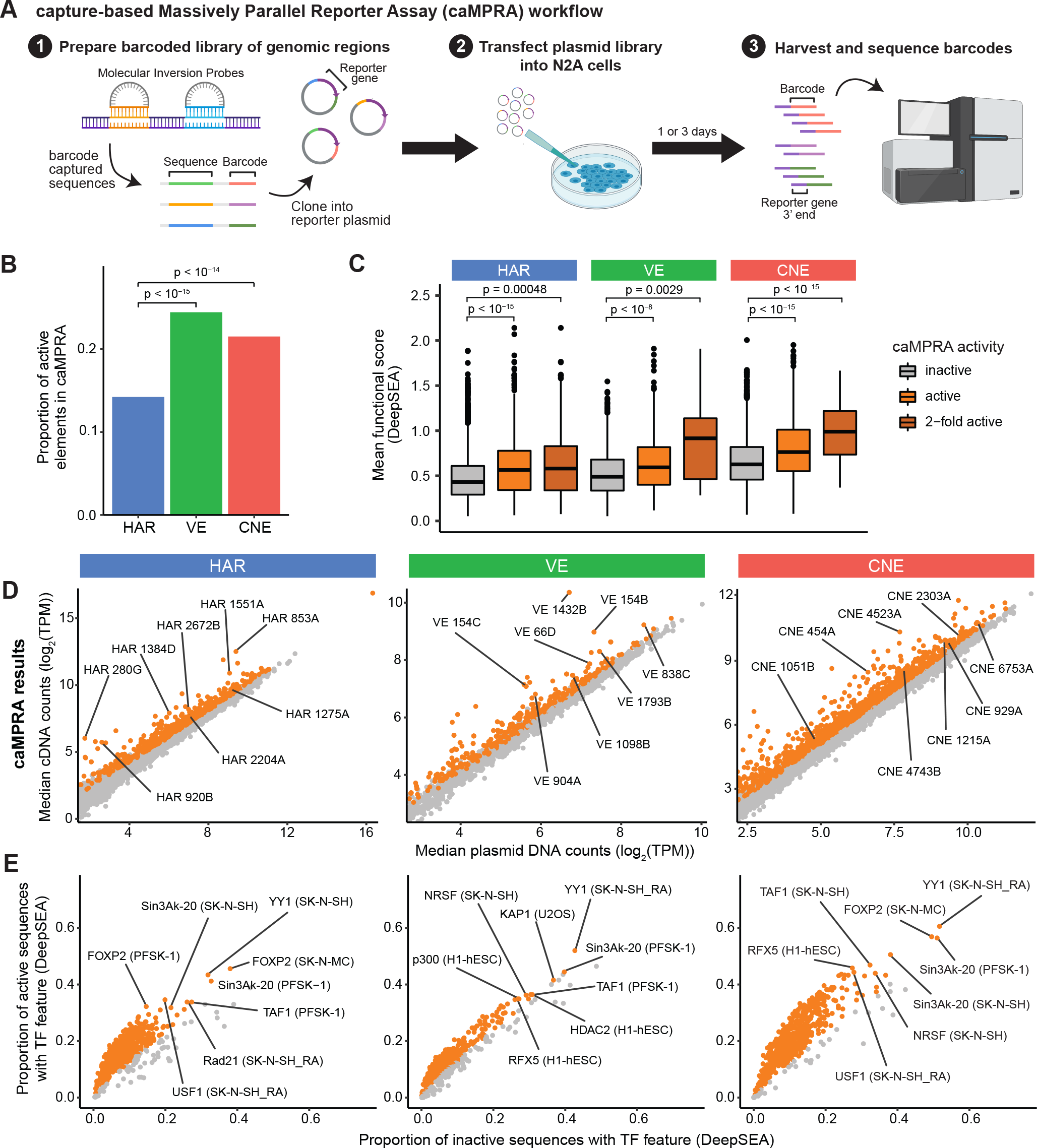
HARs, VEs, and CNEs display enhancer activity in a capture-based Massively Parallel Reporter Assay (caMPRA). (A) Schematic of caMPRA method. Sequences of interest are captured by molecular inversion probes (MIPs), barcoded, and cloned upstream of a minimal promoter driving luciferase expression. ∼500bp sequences are captured by separate MIP probes per HAR, VE, or CNE element. The enhancer reporter plasmid library is then transfected into N2A cells and cells are harvested one (D1) or three (D3) days after transfection. Transcribed barcodes and barcodes from the original plasmid library are sequenced to examine enhancer activity. The results from the D3 caMPRA experiment are shown in this figure, and the results from the D1 caMPRA experiment are shown in Fig. S6. (B) Proportion of VEs or CNEs that have enhancer activity in at least one captured sequence is significantly higher than HARs by the chi-square test after FDR correction. (C) Sequences captured from HARs, VEs, and CNEs are classified as inactive, active, or 2-fold active and compared to their predicted mean functional score from DeepSEA (average of −*log*_10_(*e− value*) for every feature) (Zhou and Troyanskaya, 2015). P-values were determined with the hypergeometric test and adjusted by FDR correction. (D) Normalized cDNA counts vs normalized plasmid counts for sequences captured from HARs, VEs, and CNEs. Sequences with significant enhancer activity are in orange. (E) TF features were predicted by DeepSEA for each captured sequence. TF features significantly enriched in active sequences by caMPRA are shown in orange. Representative TF features are marked in the format: TF (cell type).

Significantly more VEs (24.4%) and CNEs (21.5%) had enhancer activity compared to HARs (14.2%) in N2A cells (*p* < 10^−14^; Fig. 2B). This finding is consistent with the definition of VEs and CNEs as experimentally validated or predicted enchancers respectively, whereas HARs are defined solely from genomic sequence changes in the human lineage. Many of the HARs, VEs, and CNEs that have enhancer activity are located near ASD-associated genes, including HAR3091 within *IL1RAPL1*, VE2019 near *ARX*, and CNE6536 near *ASXL3* (Abrahams et al., 2013). Active elements are also enriched for the motifs of neurodevelopmental TFs including FOXP2 and TAF1 (Fig. 2E, Fig. S6E) when assessed with DeepSEA, a deep learning model trained to predict thousands of features including TF binding (Zhou et al., 2018). Although N2A cells were not among the cell lines used for training and prediction in DeepSEA, the TF enrichment predictions from DeepSEA for active elements were specific for cell lines similar to N2A cells, including the neuroblastoma cell lines SK-H-SH and SK-N-MC and the neuroectodermal cell line PFSK-1, demonstrating the specificity of our assay for the TF milieu present in these cells. Further, we observed strong concordance between caMPRA-based activity and the predicted functional score from DeepSEA for HARs, VEs, and CNEs, particularly for sequences that exhibit a 2-fold increase in enhancer activity by caMPRA (Fig. 2C, Fig. S6C).

### High-throughput mutagenesis of HARs causes gains as well as losses of enhancer activity

To determine whether single nucleotide variants in these non-coding regions could result in functional consequences in patients with ASD, we next sought to examine whether variants in these noncoding regions can affect regulatory activity. We focused on HARs and modified the caMPRA protocol to sparsely incorporate random variants into captured sequences using an error-prone PCR. Overall, we assessed 1,281 variants in 485 HARs across five replicate experiments (Materials and Methods; Fig. 3A, Fig. S8, Table S3).

**Figure 3.**
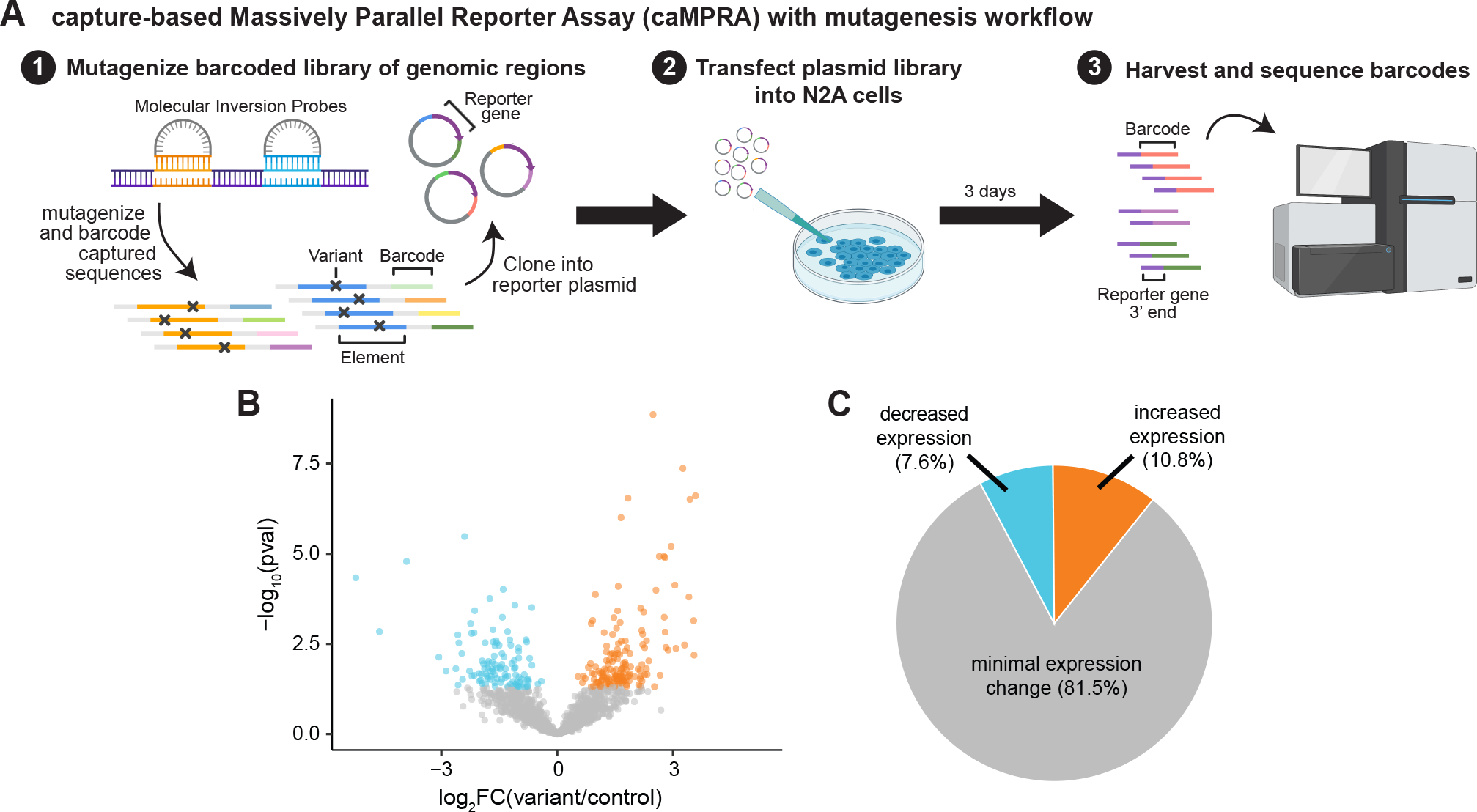
Random variants in HARs can modulate enhancer activity. (A) Schematic of caMPRA with random mutagenesis. (B) Volcano plot of fold change in expression and adjusted p-value for each mutagenized sequence. (C) Pie chart of percent of mutagenized sequences with decreased expression, increased expression, or no statistically significant change in expression.

Whereas most tested variants (81.5%) did not significantly alter regulatory activity (Fig. 3B, C), in general agreement with studies of other regulatory elements (Kircher et al., 2019; Snetkova et al., 2021), those that did were as likely to increase activity (10.8%) as to decrease it (7.6%). This is in contrast to prior mutagenesis studies where most variants that changed regulatory activity decreased expression (Kircher et al., 2019; Snetkova et al., 2021). It is possible that this difference reflects that prior studies mutagenized known enhancer elements, including VEs, whereas HARs likely comprise a mix of regulatory elements-including enhancers, repressors, and regulatory regions active in other species that have reduced or lost activity in humans. When we examine only sequences that contain a single introduced random variant, the proportion of variants that change expression and the distribution of fold changes were similar to sequences with more than 1 introduced variant (Fig. S7C, D), suggesting that single base pair changes in HARs can have profound effects on both gains and losses of enhancer activity.

### Rare, recessive variants in HARs, VEs, and CNEs are enriched in individuals with ASD in a consanguineous cohort

To examine whether HARs, VEs, and CNEs contribute to ASD risk, we examined whether there is an excess of rare, recessive variation in HARs, VEs, and CNEs in patients with ASD. Given the redundancy of regulatory networks even for highly conserved noncoding regions (Cannavò et al., 2016; Osterwalder et al., 2018), we reasoned that the bulk of our candidate enhancer sequences may act in a recessive manner, rather than via the *de novo* mode of contribution of highly constrained dominant genes. We first revisited a consanguineous cohort, the Homozygosity Mapping Collaborative for Autism (HMCA) (Morrow et al., 2008), where we had previously observed an enrichment of rare, recessive variants in HARs in ASD cases compared to controls using targeted sequencing (Doan et al., 2016). This enrichment was seen only when examining rare variants that were predicted to be damaging by conservation-based variant effect predictors (Doan et al., 2016).

When we now examine an expanded set of 3,171 HARs using whole-genome sequencing (WGS) on a larger number of families from HMCA (a total of 662 individuals including 193 probands), we continue to identify a strong enrichment of rare, recessive variants in HARs in cases compared to matched controls (*OR* = 2.142, adjusted *p* = 0.001; Fig. 4B). We defined recessive variants as variants that are homozygous, compound heterozygous, or hemizygous (specifically in male individuals for the X chromosome). Because hemizygous variants on the male X chromosome are much more likely to arise compared to homozygous variants on the female X chromosome, we only examined the autosomes when calculating combined rates for males and females, but included the X chromosome when analyzing males and females separately. As in the prior study, we observe an enrichment only when examining rare variants that are predicted to be damaging by conservation-based variant effect predictors (Materials and Methods; referred to hereafter as “conserved bases”), but not when examining non-conserved bases (adjusted *p* > 0.05).

**Figure 4.**
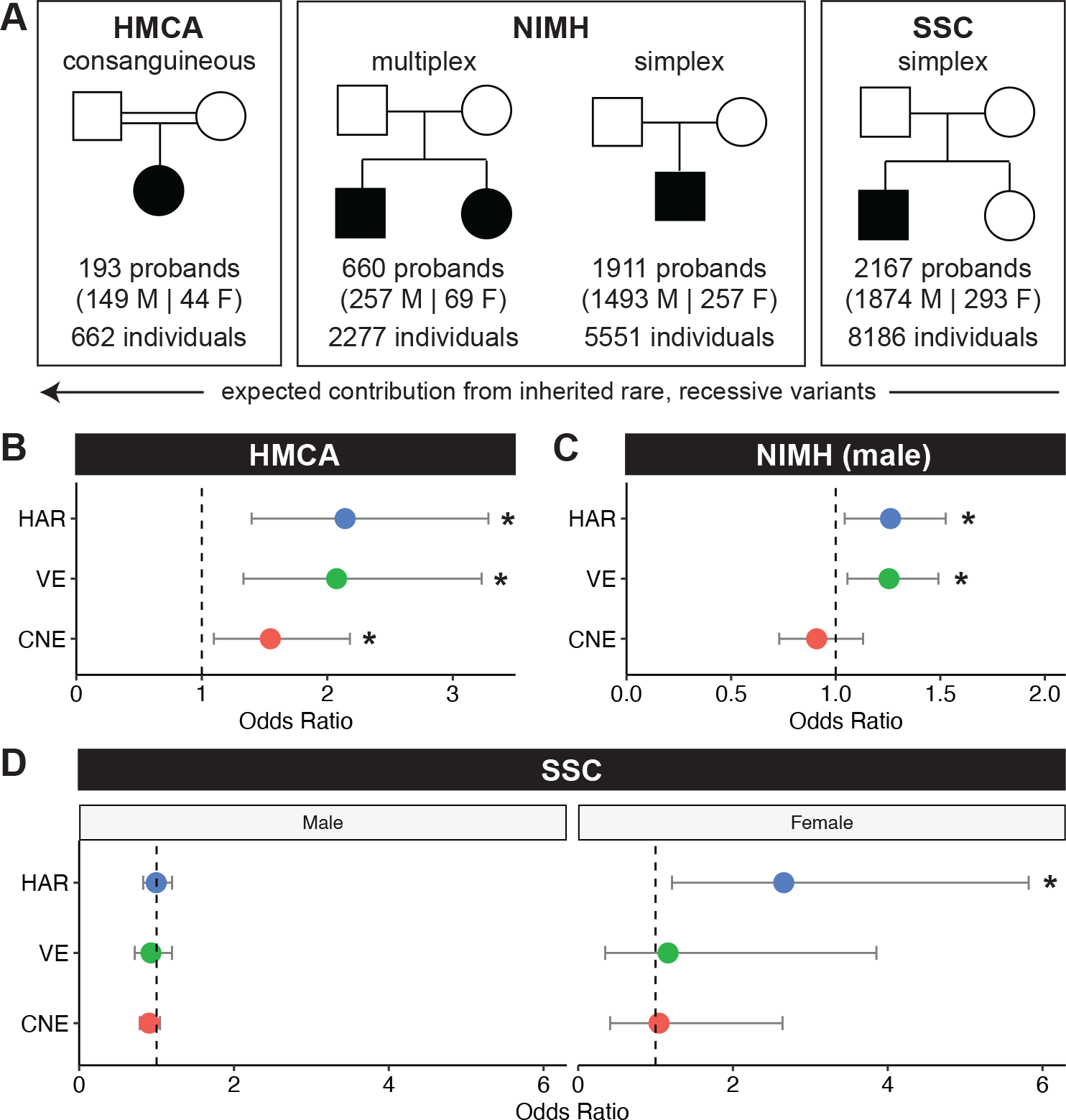
The contribution of rare, recessive variants in HARs, VEs, and CNEs to ASD varies across cohorts based on family structure. (A) We examined HARs, VEs, and CNEs in three cohorts: 1. HMCA, a consanguineous cohort; 2. NIMH, a cohort that includes multiplex and simplex families; 3. SSC, a cohort that only contains simplex families. (B) In the HMCA cohort, cases are enriched for rare, recessive variants in HARs (adjusted *p* = 0.0014), VEs (adjusted *p* = 0.0038), and CNEs (adjusted *p* = 0.0412) at allele frequency (*AF*) < 0.005. (C) In the NIMH cohort, male cases are enriched for rare, recessive variants in HARs (adjusted *p* = 0.0495) and VEs (adjusted *p* = 0.0297) at *AF* < 0.001. Combined autosomal rates for males and females were not significant, and there was insufficient sample size to assess females separately (Fig. S10A, B). (D) In the SSC cohort, female cases are enriched for rare, recessive variants in HARs (adjusted *p* = 0.0438) at *AF* < 0.005. All analyses are done on conserved bases, and allele frequency cut-offs were chosen based on predefined heuristics (Materials and Methods). Odds ratios are consistent across different *AF*s (Fig. S9A, Fig. S11, Fig. S12). Statistical analyses are detailed in Materials and Methods.

VEs also had a large excess of rare, recessive variants in conserved bases when comparing cases to controls that was similar in magnitude to the excess seen in HARs (*OR* = 2.074, adjusted *p* = 0.004; Fig. 4B), whereas CNEs had a significant, but less pronounced excess of rare, recessive variants in cases compared to controls (*OR* = 1.546, adjusted *p* = 0.041; Fig. 4B). The enrichment of rare, recessive variants in HARs, VEs, and CNEs is stable across a range of low allele frequencies, suggesting that the signal we observe is not dependent on specific allele frequency cut-offs (Fig. S9A). Although we are under-powered to assess significance when each sex is analyzed separately, similar excesses in rare, recessive variants are observed in both males and females (Fig. S9B).

The observed rates of rare, recessive variants between cases and controls (0.239 versus 0.128 for HARs, 0.216 versus 0.117 for VEs, 0.414 versus 0.313 for CNEs) yield substantial estimated contributions to ASD of 9.9%, 11.1%, and 10.0% for recessive alleles in HARs, VEs, and CNEs, respectively (Materials and Methods). Together with a previous finding in this cohort of a ∼4x excess of rare, homozygous, inherited deletions in noncoding, but not in coding, genomic regions (Schmitz-Abe et al., 2020), our results suggest that homozygous noncoding variation in this cohort contributes significantly to ASD risk by several mechanisms, and is also consistent with a relatively modest contribution of recessive exonic mutations in this cohort (Yu et al., 2013).

### Rare, recessive variants in HARs and VEs are enriched in individuals with ASD in non-consanguineous cohorts

We then examined whether the enrichment of rare, recessive variants in HARs, VEs, and CNEs is also observed in a larger, non-consanguineous cohort from the NIMH repository. We expect effect sizes for recessive variants to be considerably smaller in non-consanguineous cohorts compared to consanguineous cohorts where both direct consanguinity and endogamy make it more likely that the same rare variant is inherited from both parents (Bittles and Black, 2010). However, compared to the slightly less than 200 probands in the consanguineous cohort, the NIMH repository contains >2000 affected probands, offering greater resolution to detect small differences in recessive variants and to identify a larger set of patient variants for functional studies (Fig. 4A). We examined 660 probands from multiplex families, where inherited variants are more likely to play a role (Ruzzo et al., 2019; Cirnigliaro et al., 2023), and 1911 probands from families with only 1 affected child (either with or without an unaffected sibling). The latter are likely to be simplex families, where recessive variants have a lower contribution to disease compared to *de novo* mechanisms (Sebat et al., 2007; Marshall et al., 2008).

Targeted sequencing of HARs, VEs, and CNEs with molecular inversion probes (Materials and Methods) showed a non-significant excess of rare, recessive variants in HARs and VEs at conserved bases when considering males and females jointly (HARs: *OR* = 1.196, adjusted *p* = 0.186; VEs: *OR* = 1.193, adjusted *p* = 0.069; Fig. S10A), while males considered alone, which captures hemizygous variants on chromosome X, revealed significant enrichment for rare, recessive variants in both HARs and VEs at conserved, but not at less conserved, bases (HARs: *OR* = 1.262, adjusted *p* = 0.050; VEs: *OR* = 1.255, adjusted *p* = 0.030; Fig. 4C) across allele frequency cut-offs (Fig. S11). In contrast, CNEs were not enriched for rare, recessive variants in cases versus controls for males (males: *OR* = 0.909, adjusted *p* = 1; Fig. 4C). Because females are much less likely than males to be diagnosed with ASD (Robinson et al., 2013), we have a much smaller number of female individuals in this cohort and were under-powered to examine females alone given the odds ratios observed in males (Fig. S10B). Notably, the odds ratios are similar when comparing males and females jointly or males separately. Consistent with the effect of family structure on the contribution of recessive variants to ASD risk, we also observe a larger effect size in multiplex families (HARs: *OR* = 1.645, VEs: *OR* = 1.378) compared with the likely simplex families (HARs: *OR* = 1.189, VEs: *OR* = 1.180) in males (Fig. S10C).

The rates of rare, recessive variants between male cases and controls are 0.141 versus 0.115 for HARs and 0.213 versus 0.181 for VEs, resulting in an estimated contribution of recessive alleles in HARs and VEs to 2.6% and 3.7% of ASD cases respectively. This contribution is similar to the 3-5% contribution of rare, recessive coding variants to ASD cases in a similar cohort (Doan et al., 2019), but smaller than the estimated ∼10% contribution of rare, recessive alleles in HARs, VEs, and CNEs to ASD cases we observed in the consanguineous cohort. These results highlight the unique suitability of consanguineous cohorts to examine noncoding variation, but nevertheless confirm the contribution of HARs and VEs to ASD and provide a large set of rare, potentially damaging variants for further study (Table 1, Table S4).

**Table 1:** Examples of HARs, VEs, and CNEs that have more variants found in cases compared to controls. Full list in Table S4. Asterisks indicate genes that are loss-of-function intolerant (pLI > 0.9) (Lek et al., 2016). Potential target genes were determined by gene proximity and by location within the same topologically associated domain (Dixon et al., 2012, 2015). For HAR3162, one variant was observed in both a case and a control individual in HMCA and was excluded from the table. Coordinates are in hg19. ID: intellectual disability. DD: developmental disorders.

| Element | Cohort | Variants found in cases and not controls (hg38) | Number of cases with variants | Number of controls with variants | Potential target genes | Disease and functional associations |
| --- | --- | --- | --- | --- | --- | --- |
| HAR1362 | NIMH | chr2:44493977 (G->A)<br>chr2:44494211 (G->A) | 2 | 0 | *CAMKMT*, *SIX3* *, *PREPL* | Required for development of anterior neural structures (*SIX3*) (Lagutin et al., 2003) |
| HAR1479 | NIMH | chr2:145221015 (G->A)<br>chr2:145221025 (C->A) | 2 | 0 | *ZEB2* *, *GTDC1*, *ARHGAP15* | Mutations cause Mowat-Wilson syndrome (*ZEB2*) (Epifanova et al., 2019) |
| HAR3094 | NIMH | chrX:30371544 (G->A)<br>chrX:30371553 (A->G) | 2 | 0 | *NR0B1* *, *CXorf21*, *IL1RAPL1* *, *MAGEB1*, *MAGEB2*, *MAGEB3* | Mutations associated with ASD and ID (*IL1RAPL1*) (Bhat et al., 2008; Mikhail et al., 2011) |
| HAR3134 | HMCA, NIMH | chrX:122662679 (T->C)<br>chrX:122662692 (A->G) | 3 | 0 | *GRIA3* * | Mutations associated with ASD, X-linked syndromic ID, and schizophrenia (*GRIA3*) (Wu et al., 2007; Guilmatre et al., 2009; Singh et al., 2022) |
| HAR3162 | NIMH | chrX:144625836 (G->C)<br>chrX:144625865 (A->G)<br>chrX:144625878 (G->A)<br>chrX:144625958 (G->A) | 4 | 1 | *SLITRK2*, *SLITRK4* | Mutations associated with ID, DD, and neuropsychiatric symptoms (*SLITRK2*) (El Chehadeh et al., 2022) |
| VE15 | NIMH | chr1:10737260 (T->C)<br>chr1:10737343 (G->C) | 2 | 0 | *CASZ1* * | Mutations associated with ASD, ID, and DD (*CASZ1*) (Coe et al., 2019) |
| VE162 | NIMH | chr1:213425381 (T->C)<br>chr1:213425533 (A->C) | 2 | 0 | *PROX1* *, *RPS6KC1*, *SMYD2* | Regulates interneuron differentiation (*PROX1*) (Miyoshi et al., 2015) |
| VE235 | NIMH | chr2:63049151 (C->A) | 1 | 0 | *OTX1* * | Mutations associated with ASD (Liu et al., 2011) |
| VE462 | NIMH | chr3:147847042 (T->C)<br>chr3:147847133 (T->C)<br>chr3:147847216 (T->C) | 3 | 0 | *ZIC1* *, *ZIC4* | Involved in medial telencephalon development (*ZIC1*) (Inoue et al., 2007) |
| VE644 | NIMH | chr5:88396771 (G->T)<br>chr5:88397035 (C->T) | 2 | 0 | *MEF2C* *, *TMEM161B* | Mutations associated with ASD (*MEF2C*) (Novara et al., 2010); Mutations associated with polymicrogyria (*TMEM161B*) (Akula et al., 2023; Wang et al., 2023) |
| CNE6445 | HMCA | chr17:69607668 (C->T) | 2 | 0 | *KCNJ16*, *MAP2K6* *, *KCNJ2* | Member of MAP/ERK pathway, which has been linked to changes in social behavior (*MAP2K6*) (Albert-Gasco et al., 2020) |
| CNE7200 | HMCA | chrX:18424091 (T->C) | 2 | 0 | *CDKL5* * | Mutations associated with Rett syndrome and epilepsy (*CDKL5*) (Weaving et al., 2004) |

We next examined the Simon Simplex Collection (SSC), which consists of 8,186 individuals with WGS data and is specifically limited to simplex families with a single proband and unaffected siblings (Fischbach and Lord, 2010) (Fig. 4A). In such a cohort, recessive effects are expected to be minor, and potentially undetectable, compared to the consanguineous HMCA cohort and the NIMH cohort that contained multiplex families (Sebat et al., 2007; Yu et al., 2013). Indeed, we observed no excess of rare, recessive variants in HARs, VEs, or CNEs in almost all comparisons. When examining the cohort separated by sex, we do find a significant excess of rare, recessive variants in HARs in female ASD cases in conserved, but not at less conserved, bases (*OR* = 2.657, adjusted *p* = 0.044; rate of rare, recessive variants is 0.027 in cases and 0.010 in controls for an estimated 1.7% contribution; Fig. 4D) across allele frequency cut-offs (Fig. S12), with no similar enrichment in males, despite there being 1874 male probands and only 293 female probands in SSC. An enrichment in females, but not in males, may reflect the female protective effect, a phenomenon where female probands require a higher genetic burden (potentially including variants in HARs) than male probands to develop ASD (Gilman et al., 2011; Robinson et al., 2013; Jacquemont et al., 2014), and also parallels the larger contribution to ASD of recessive coding variants in females compared to males (Doan et al., 2019).

Our findings from multiple ASD cohorts indicate that rare, recessive variants in HARs, VEs, and CNEs contribute to ASD burden. As expected, the impact of these variants tracks the known contribution of recessive variants as a function of family structure, with by far the largest contribution seen in consanguineous families, a smaller contribution seen in male ASD cases in a cohort of non-consanguineous families that includes multiplex families, and a contribution that is only discernible in females among simplex families. Consanguineous families have been used to uncover noncoding variants underlying risk for other diseases (Schultz et al., 2009; Bae et al., 2014; Favaro et al., 2014; Tuncay et al., 2022), and our results suggest that further expanding consanguineous cohorts may be particularly suitable for identifying and analyzing noncoding contributions to disease.

### Variants enriched in ASD patients implicate new genes in ASD risk

While we are underpowered to pinpoint specific HARs, VEs, or CNEs that are statistically enriched for patient variants, individual HARs, VEs, or CNEs with a numerical excess of rare, recessive variants in cases compared to controls represent potential candidates for further study, particularly since the number of controls far exceeds the number of cases in each cohort. We focused on rare, recessive variants enriched in ASD cases compared to controls in HARs, VEs, or CNEs from the HMCA cohort and in HARs or VEs from the NIMH cohort because there is a greater expected contribution of inherited variants from those cohorts compared to the simplex SSC cohort (Sebat et al., 2007; Bittles and Black, 2010; Yu et al., 2013).

HARs, VEs, and CNEs enriched for variants found in cases compared to controls are located near both ASD-associated genes and genes that have not been previously linked to ASD. Intriguingly, proteins encoded by many of the newly identified candidate genes are known to interact with proteins encoded by ASD-associated genes (Fig. S13A). This reflects recent studies that identify convergent effects on protein networks across multiple, distinct genetic models of ASD (Paulsen et al., 2022; Pintacuda et al., 2023; Li et al., 2023). Many newly identified candidate genes, as well as many genes previously associated with ASD, are also loss-of-function intolerant (blue circles indicate genes with pLI > 0.9 in Fig. S13A, B), suggesting that expression changes in specific cell types (such as through regulatory variants in HARs, VEs, or CNEs) may have significant phenotypic consequences.

We highlight a subset of HARs, VEs, and CNEs that are enriched for patient variants in Table 1 (full list in Table S4). These include HAR3094 and VE644, which are near the ASD-associated genes *IL1RAPL1* (Bhat et al., 2008; Mikhail et al., 2011; Montani et al., 2019) and *MEF2C* (Novara et al., 2010) respectively. In contrast, HAR3162 (near *SLITRK2*), VE162 (near *PROX1*), and CNE6445 (near *MAP2K6*) are located near promising candidate genes that have not been previously associated with ASD. Mutations in *SLITRK2* result in moderate to severe intellectual disability with a range of behavioral and neuropsychiatric symptoms (El Chehadeh et al., 2022). In E11.5 embryonic mice, we find that HAR3162 has enhancer activity in the ventral telencephalon (Fig. S13C, D), where *SLITRK2* is expressed (Magdaleno et al., 2006; Diez-Roux et al., 2011), suggesting that HAR3162 may regulate *SLITRK2* expression. Similarly, VE162 has enhancer activity in the ventral telencephalon in E11.5 embryonic mice (Visel et al., 2007) and has been shown to physically interact with the promoter of *PROX1* (Song et al., 2020), a gene that regulates interneuron differentiation in the ventral telencephalon (Magdaleno et al., 2006; Miyoshi et al., 2015). Finally, CNE6445 is located near the loss-of-function intolerant MAP kinase gene *MAP2K6*. MAP/ERK signaling regulates many aspects of normal brain development, and dysregulation of this pathway has been associated with changes to social behaviors (Albert-Gascó et al., 2020).

### HARs enriched for ASD patient variants regulate the neurodevelopmental gene IL1RAPL1

As an initial functional investigation into whether HARs, VEs, or CNEs that are enriched for patient variants might contribute to ASD risk, we characterized HAR3091 and HAR3094 (Fig. 5A). Both of these HARs reside near the gene *IL1RAPL1*, act as enhancers by caMPRA, and are enriched for patient variants. In the NIMH cohort, HAR3091 and HAR3094 each contained two variants in cases and none in controls at conserved bases (Table 1, Table S4).

**Figure 5.**
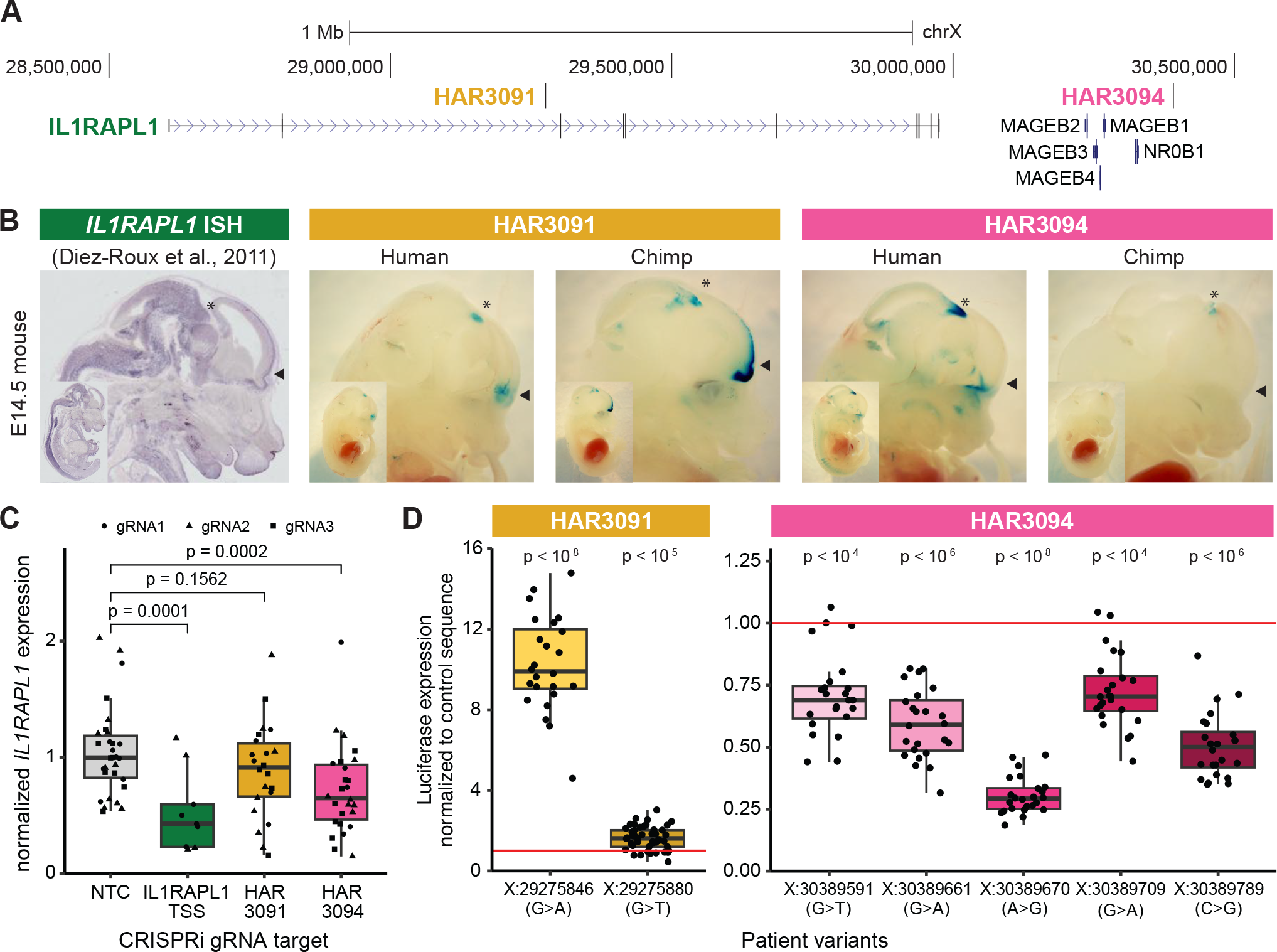
Patient variants in HAR3091 and HAR3094 likely regulate *IL1RAPL1* expression in multiple brain regions. (A) The genomic region containing *IL1RAPL1*, HAR3091, and HAR3094. (B) Constructs containing either the human or chimpanzee version of HAR3091 and HAR3094 cloned upstream of a minimal promoter driving lacZ expression were randomly integrated into mice and analyzed at E14.5. HAR3091 has enhancer activity predominantly in the telencephalon and olfactory bulb (filled arrowheads), and the human version of HAR3091 is a weaker enhancer than the chimpanzee version. In contrast, HAR3094 has enhancer activity predominantly in the midbrain (asterisks), and the human version of HAR3094 is a stronger enhancer than the chimpanzee version. Representative embryos are shown (all embryos are in Fig. S14). *In situ* hybridization of *IL1RAPL1* at E14.5 from the Eurexpress database (Diez-Roux et al., 2011) is shown for comparison. (C) CRISPRi targeting the *IL1RAPL1* TSS and HAR3094 significantly decrease *IL1RAPL1* expression compared to the non-targeting control (NTC) gRNAs in iPSC-derived neurons induced by *NGN2* expression. Multiple gRNAs were tested per target region. (D) Patient variants in HAR3091 and HAR3094 were tested for luciferase expression in N2A cells. HAR3091 patient variants significantly increased luciferase expression, whereas HAR3094 patient variants significantly decreased luciferase expression. Statistical analyses are detailed in Materials and Methods. Coordinates are in hg19.

*IL1RAPL1* is a gene important for synaptic density and dendrite formation at excitatory synapses (Montani et al., 2019) that is highly loss-of-function intolerant (Lek et al., 2016; Karczewski et al., 2020). Exonic point mutations, deletions, and duplications of *IL1RAPL1* have been associated with ASD and intellectual disability (Carrié et al., 1999; Tabolacci et al., 2006; Froyen et al., 2007; Piton et al., 2008; Firth et al., 2009; Behnecke et al., 2011; Franek et al., 2011; Du et al., 2018; Montani et al., 2019), suggesting that *IL1RAPL1* is dosage-sensitive to both gain and loss of expression. HAR3091 is located in the second intron of *IL1RAPL1*, and HAR3094 is located downstream of *IL1RAPL1* within the same topologically associated domain (Won et al., 2016).

Transgenic mouse analysis suggests that HAR3091 and HAR3094 have regionally restricted and species-specific enhancer activity in the developing brain. We generated transient transgenic mice with either the human or chimpanzee versions of HAR3091 or HAR3094 located upstream of a minimal promoter driving a *lacZ* reporter gene (Fig. 5B, Fig. S14). We harvested mouse embryos at E14.5, when *in situ* data show strong *IL1RAPL1* expression (Diez-Roux et al., 2011), and stained for lacZ expression (Materials and Methods). HAR3091 drives lacZ expression predominantly in the telencephalon and olfactory bulb (arrowheads in Fig. 5B and Fig. S14), with more limited lacZ expression in the midbrain (asterisks in Fig. 5B and Fig. S14). In contrast, HAR3094 drives lacZ expression predominantly in the midbrain, with more limited expression in the telencephalon and the olfactory bulb. Interestingly, the chimpanzee version of HAR3091 drives much more robust activity in the telencephalon compared to the human version of HAR3091, whereas the human version of HAR3094 drives much more robust activity in the midbrain than the chimpanzee version of HAR3094. These results suggest that HAR3091 is primarily a telencephalon enhancer that has decreased activity in humans compared to chimpanzees, whereas HAR3094 is primarily a midbrain enhancer that has increased activity in humans compared to chimpanzees. Both HAR3091 and HAR3094 enhancer domains overlap with regions where *IL1RAPL1* is expressed at E14.5 (Diez-Roux et al., 2011) (Fig. 5B).

To directly test whether HAR3091 and HAR3094 might regulate *IL1RAPL1* expression, we used CRISPR inhibition (CRISPRi), which uses a nuclease-inactive Cas9 variant tethered to a KRAB domain (dCas9-KRAB) to heterochromatize and silence the target region (Gilbert et al., 2014). We induced *NGN2* expression (Materials and Methods) to differentiate human iPSCs into a heterogenous mixture of excitatory neurons that resemble neurons derived from multiple brain regions, including the regions where HAR3091 and HAR3094 have enhancer activity (Lin et al., 2021). Whereas targeting the *IL1RAPL1* TSS significantly decreased *IL1RAPL1* expression compared to NTC gRNAs as expected (adjusted *p* = 0.0001; Fig. 5C), we also observed a significant decrease in *IL1RAPL1* expression when targeting HAR3094 (adjusted *p* = 0.0002), suggesting that HAR3094 acts as an *IL1RAPL1* enhancer. When targeting HAR3091, the median *IL1RAPL1* expression decreased nominally by 8.8% but did not reach statistical significance. Given that human HAR3091 acts as a weak enhancer in transgenic mice, it is possible that our CRISPRi assay lacked the required sensitivity to detect a significant decrease in expression, especially given the wide variability in gRNA efficacy (Fig. S15). These results suggest that HAR3094, and possibly also HAR3091, are *IL1RAPL1* enhancers in neurons, and that HAR3094 is a stronger enhancer than HAR3091.

Next, we asked whether patient variants may affect the enhancer activity of HAR3091 and HAR3094. Based on availability of patient DNA, we examined one of the two rare, recessive patient variants at conserved bases in HAR3091 and the two rare, recessive patient variants at conserved bases in HAR3094. In addition, we also examined an additional patient variant in HAR3091 and three additional patient variants in HAR3094 that are rare, recessive variants but at less conserved bases. HAR3091 or HAR3094 sequences containing these variants were cloned upstream of a minimal promoter driving luciferase expression, and luciferase activity was assessed in N2A cells (Materials and Methods). Strikingly, we find that patient variants for HAR3091 significantly increased luciferase activity compared to the control HAR3091 sequence, and that patient variants for HAR3094 significantly decreased luciferase activity compared to the control HAR3094 sequence (Fig. 5D, Fig. S16). The largest effect sizes were observed for the patient variants at conserved bases, consistent with the established link between conservation and functional activity and our finding that an excess of rare, recessive variants is observed in ASD cases compared to controls for conserved but not less conserved bases. These results indicate that patient variants modulate HAR3091 and HAR3094 enhancer activity and may result in changes to *IL1RAPL1* expression in specific brain regions.

### ASD patient variants near SIM1, a human neurobehavioral gene, modulate in vivo enhancer activity

VE854, commonly referred to as hs576 (Visel et al., 2007), is an enhancer of the nearby obesity-associated gene *SIM1* (Ahituv et al., 2007; Bonnefond et al., 2013; Kim et al., 2014; Matharu et al., 2019) and contains two rare, recessive patient variants in the HMCA and NIMH cohorts (Fig. 6, Table S4). *SIM1* loss-of-function has been associated with obesity and neurobehavioral deficits; in one study that identified 13 obese individuals with rare, *de novo SIM1* mutations, 11 also presented with neurobehavioral abnormalities including ASD (Ramachandrappa et al., 2013). Genes downstream of *SIM1* have similarly been associated with both obesity and neurological phenotypes (Kasher et al., 2016), suggesting that modulating this pathway may contribute to obesity, ASD, and their comorbidity.

**Figure 6.**
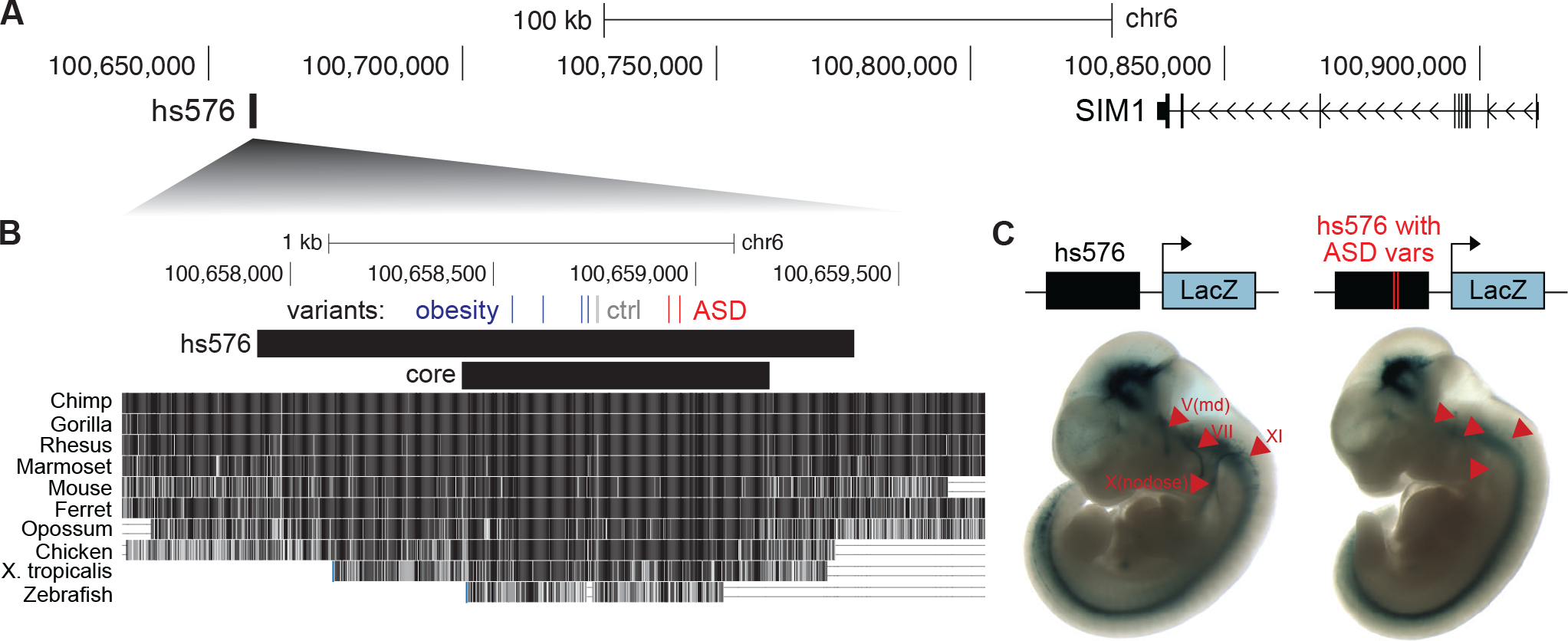
Patient variants in hs576 (VE854) reduce enhancer activity in cranial nerves. (A) The genomic region containing hs576 and *SIM1*. (B) The locations of two ASD patient variants from the HMCA and NIMH cohorts (ASD; red), two variants from one control individual (ctrl; gray), and four previously identified obesity-associated variants (blue) (Kim et al., 2014) are indicated. These variants are all located in the most conserved region of hs576 (core), which recapitulates most of the enhancer activity of the entire hs576 element (Kim et al., 2014). (C) Constructs containing hs576 without (*n* = 4) or with (*n* = 6) the two ASD patient variants upstream of a minimal promoter driving the lacZ gene were integrated into the safe-harbor H11 locus and analyzed for lacZ expression at E11.5 (Materials and Methods). Arrowheads indicate cranial nerves where the inclusion of the two ASD patient variants reduces enhancer activity. Representative embryos are shown (all embryos are in Fig. S17). Coordinates are in hg19.

Hs576 has been previously found to drive enhancer activity in the developing brain, somites, and cranial nerves in transgenic E11.5 mice and in the forebrain and hippocampus in E14.5 mice, matching the expression pattern of *SIM1* (Visel et al., 2007; Kim et al., 2014). This enhancer activity is mainly derived from the most highly conserved portion of hs576 (“core” in Fig. 6B) (Kim et al., 2014). Intriguingly, rare variants associated with obesity (Kim et al., 2014) and our identified ASD patient variants are both located in this core region, albeit in separate clusters at the 5’ and 3’ ends respectively (Fig. 6B). There is also one control individual in the NIMH cohort that contains two neighboring variants in hs576 (Fig. 6B). To test whether the ASD patient variants affect the enhancer activity of hs576, we first confirmed its known expression pattern in E11.5 mice by integrating a construct containing hs576 upstream of a minimal promoter driving lacZ expression at the H11 safe-harbor locus (Materials and Methods; Fig. 6C). We then generated E11.5 transgenic mice where hs576 containing the two ASD patient variants was integrated at the H11 locus. Strikingly, we find that hs576 containing the two ASD patient variants had reduced or absent enhancer activity in multiple cranial nerves, in particular the mandibular portion of the trigeminal nerve (V), the facial nerve (VII), the nodose nerve / inferior part of vagus nerve (X), and the accessory nerve (XI) across multiple embryos (arrowheads in Fig. 6C; Fig. S17). These results suggest that ASD patient variants can alter *SIM1* expression and may contribute to ASD.

## Discussion

In this study, we identify HARs, VEs, and CNEs as three classes of noncoding regions that are near ASD-associated and dosage-sensitive genes, and show contrasting degrees of evolutionary conservation, constraint within humans, and predicted functional activity in the brain. Using caMPRA, we find that VEs and CNEs are significantly more likely than HARs to act as enhancers in N2A cells, and that single nucleotide variants in HARs can both increase and decrease enhancer activity. We identify a substantial contribution of rare, recessive variation in these noncoding elements to ASD risk but one that varies sharply with family structure: In a consanguineous cohort, where pathogenic *de novo* mutations are relatively less common compared to pathogenic inherited variants, recessive variation in HARs, VEs, and CNEs each contributes to ∼10% of ASD cases. This is a substantial contribution when compared to the ∼13% contribution of homozygous, noncoding copy number variants (Schmitz-Abe et al., 2020) and the ∼30% contribution of homozygous, loss-of-function and missense variants (Yu et al., 2013; Doan et al., 2019) to ASD cases previously identified in this consanguineous cohort. In a non-consanguineous cohort of simplex and multiplex families, we observe a smaller (∼ 3%) contribution of recessive variation in HARs and VEs to male ASD cases, comparable to the 3-5% contribution of recessive coding variants observed in a similar cohort (Doan et al., 2019). In contrast, the contribution of recessive variation in these noncoding elements to ASD risk is almost completely absent in an entirely simplex cohort, where *de novo*, rather than inherited, variants are more likely to contribute to ASD risk (Sebat et al., 2007; Yu et al., 2013; Cirnigliaro et al., 2023).

It is somewhat surprising that HARs consistently have the highest odds ratios for rare, recessive variants in ASD compared to controls across all three cohorts, followed by VEs and then by CNEs. Given that VEs are experimentally validated as neural enhancers and CNEs show strong predictions of neural enhancer activity, one might expect that VEs and CNEs would have a stronger contribution to ASD risk than HARs, which are defined solely by brain-agnostic signatures of selection. The strong enrichment we observe in HARs may suggest that regions that are recently evolved in humans are more likely to contribute to disease risk than conserved regions. Intriguingly, two new sets of HARs were identified after the completion of this study (Keough et al., 2023; Bi et al., 2023). Although we are not powered to statistically assess these new, smaller HAR sets, both sets are also nominally enriched for rare, recessive variants in ASD cases compared to controls in the HMCA cohort (Fig. S18). Our results extend recent findings that common variation in noncoding, human-evolved regions may contribute to risk for neurological diseases (Xu et al., 2015; Srinivasan et al., 2016; Song et al., 2018), and suggests that rare variation in human-evolved regions may also preferentially contribute to human disease risk.

The proportion of HARs that showed enhancer activity in the caMPRA experiment was significantly lower than VEs and CNEs (Fig. 1D, Fig. 2B), which may reflect that HARs include regions with different regulatory functions. For instance, HARs include regions such as HAR3091 that were previously strong enhancers in other mammals, but where decreasing or silencing enhancer function appears to have been selected for in the human lineage. Variability in whether human-specific sequence variants in HARs increase or decrease enhancer activity has been observed in multiple MPRAs comparing human and chimpanzee HAR sequences (Uebbing et al., 2021; Girskis et al., 2021; Whalen et al., 2023). We also clearly observe this heterogeneity in HAR function in the caMPRA mutagenesis experiment where a similar proportion of single nucleotide variants increase and decrease enhancer activity in HARs (Fig. 3C). In contrast, prior mutagenesis studies that examined known enhancers found that most functional variants decreased enhancer activity (Kircher et al., 2019; Snetkova et al., 2021). The strong enrichment for rare, recessive variants in HARs suggests the importance of examining noncoding regions that perform different regulatory functions for their contributions to ASD risk. In contrast to HARs, CNEs had the lowest odds ratios for recessive ASD risk in all cohorts, despite being more highly conserved across species and more highly constrained within humans than HARs and VEs. In addition to a high level of conservation and constraint, CNEs are also predicted by the Roadmap Epigenomics Project to be active in a large number of body tissues outside of the brain (Fig. S1). This raises the possibility that variants in CNEs may be more likely to have large effect sizes or lead to pleiotropic effects that act in a dominant, *de novo* fashion rather than in a recessive, inherited fashion. Although we do not observe a case-specific enrichment of *de novo* variants in CNEs in the SSC cohort (Fig. S19), it is possible that an increased sample size may reveal a contribution of *de novo* variants in CNEs to ASD risk.

Prior work in non-consanguineous multiplex cohorts did not detect a significant contribution of noncoding, inherited variation when examining regions predicted to be functional (using similar heuristics as we use here to define CNEs) and suggested that sample sizes of 8000-9000 probands would be required for sufficient statistical power (Cirnigliaro et al., 2023). In contrast, we find a significant enrichment for rare, recessive variants for HARs, VEs, and CNEs in a consanguineous cohort with only 193 probands and confirm this enrichment for HARs and VEs in a multiplex cohort. This suggests (1) that HARs and VEs, which are less likely than CNEs to be predicted to be active by epigenomic data (Fig. 1) but which were defined either by evolutionary signatures of selection in humans or by experimentally validated enhancer activity, are particularly impactful sets of noncoding regions, and that current predictors of functional activity require improvement; and (2) that consanguineous families may be especially suitable for analyzing noncoding contributions to disease risk, given that both direct consanguinity and endogamy enhance potential recessive contributions (Bittles and Black, 2010).

Although our cohorts are too small to ask whether individual noncoding elements are statistically enriched for rare, recessive variants, we posit that noncoding elements that show a numerical excess of rare, recessive variants in ASD cases compared to controls are nevertheless promising candidates for further study. We find that genes near noncoding elements with an excess of rare, recessive variants in cases compared to controls include both known ASD-associated genes and previously unassociated genes. Strikingly, proteins encoded by both ASD-associated and previously unassociated genes are known to interact, mirroring recent studies that identify convergent effects on protein networks across multiple, distinct genetic models of ASD (Paulsen et al., 2022; Pintacuda et al., 2023; Li et al., 2023). Many of these previously unassociated genes are dosage-sensitive, known to play critical roles in neurodevelopment, or mutated in severe developmental disorders (Table 1, Fig. S13). This suggests a model whereby coding variants in these genes lead to embryonic lethality or to multi-system developmental disorders, but noncoding variants in nearby regulatory sequences dysregulate gene expression in specific cell types or at specific developmental timepoints to contribute to ASD risk.

We closely investigated two HARs, HAR3091 and HAR3094, located near *IL1RAPL1* that contain rare, recessive variants in ASD cases but not in controls. We find that these HARs modulate *IL1RAPL1* expression in distinct brain regions; HAR3091 is a weak forebrain enhancer whereas HAR3094 is a strong midbrain enhancer in humans. We further find that ASD patient variants increase enhancer activity in HAR3091, but decrease enhancer activity in HAR3094. For both HAR3091 and HAR3094, patient variants reverted enhancer activity to more closely match the strength of the chimpanzee (ancestral) sequence. Together with studies showing that disease-associated variants in other human-evolved regions move functional activity in the derived direction (Huang et al., 2022), this suggests that disease variants in human-evolved regions may act either by reverting functional activity toward an ancestral state or by pushing functional activity toward an even more derived state. The opposing direction-of-effects of HAR3091 and HAR3094 patient variants further highlight that patient variants in noncoding regions may be more likely to act on dosage-sensitive genes like *IL1RAPL1*, where moderate increases or decreases in gene expression may result in phenotypic consequences.

*IL1RAPL1* encodes a post-synaptic organizer of excitatory synapses that has been repeatedly implicated in ASD and intellectual disability both by loss-of-function as well as by duplication (Carrié et al., 1999; Tabolacci et al., 2006; Froyen et al., 2007; Piton et al., 2008; Firth et al., 2009; Behnecke et al., 2011; Franek et al., 2011; Du et al., 2018; Montani et al., 2019). *IL1RAPL1* knock-out mice have deficits in learning and memory, reduced dendritic spine density, and a reduced number of excitatory synapses (Pavlowsky et al., 2010; Houbaert et al., 2013; Yasumura et al., 2014; Montani et al., 2017), whereas over-expression of *IL1RAPL1* in cultured neurons leads to an increased number of excitatory synapses (Pavlowsky et al., 2010; Valnegri et al., 2011). Changing excitatory synapses without affecting inhibitory synapses may result in an excitatory-inhibitory synaptic imbalance, which is one of the hallmark cellular phenotypes observed in ASD models (Nelson and Valakh, 2015). Additional studies will be needed to fully understand the role of *IL1RAPL1* and how modulation of its expression by HAR3091 and HAR3094 in specific brain regions may impact ASD risk. Nevertheless, our findings linking patient variants in HAR3091 and HAR3094 to *IL1RAPL1* expression suggest that noncoding regions that contain a numerical excess of rare, recessive variants are promising candidates for functional study.

We also investigated patient variants in hs576 (VE854), an enhancer of *SIM1*, which has been associated with both obesity and neurobehavioral deficits (Ramachandrappa et al., 2013), and found that patient variants reduced enhancer activity in multiple cranial nerves. Recent research has increasingly linked peripheral nervous system deficits, such as autonomic system or sensory neuron dysregulation, to common ASD symptoms, including decreased social engagement, flat facial expressions and intonation, touch and taste sensitivity, and gastrointestinal issues (Orefice et al., 2016; Jin and Kong, 2017; Huzard et al., 2022). These symptoms are likely driven, at least in part, by cranial nerves, including the trigeminal, facial, and vagus nerves affected by the patient variants in hs576. The vagus nerve, in particular, is also important in appetite regulation (de Lartigue, 2016), and its dysregulation in a subset of individuals with ASD may underlie the comorbidity of obesity and ASD (Phillips et al., 2014). Although both the previously identified obesity-associated variants and our newly identified ASD patient variants are located in the core enhancer region of hs576, detailed weight information for the ASD patients containing variants in hs576 is not available, and future research will be needed to determine whether these ASD patient variants affect *SIM1* expression in ways that solely contribute to neurobehavioral deficits or that may also contribute to obesity. Collectively, these findings identify classes of noncoding regions that contribute to ASD disease risk and nominate specific noncoding elements and ASD patient variants for future study. We find that both regions that are highly conserved across species and likely act as neural enhancers (VEs and CNEs) and regions that are under positive selection in the human lineage and perform heterogenous regulatory functions (HARs) contribute to ASD disease risk. This highlights the importance of examining a diverse set of noncoding regions for their contribution to disease risk, including human-evolved elements and noncoding regions with diverse regulatory functions. Further, our data also demonstrate the importance of expanding cohort enrollment to diverse populations, and potentially focusing on populations with high rates of consanguinity and endogamy, since such families may be very powerful for elucidating the contribution of noncoding regions to ASD and other diseases.

## Data Availability

All data produced in the present study are available upon reasonable request to the authors

## Acknowledgements

We thank members of the Walsh and Doan labs for useful discussions. C.A.W. and R.N.D. are supported by the Allen Discovery Center program, a Paul G. Allen Frontiers Group advised program of the Paul G. Allen Family Foundation, and the Tan Yang Autism Center at Harvard University. T.S. is supported by the Stuart H.Q. & Victoria Quan Fellowship. J.H.T.S. is supported by the Y. Eva Tan Fellowship from the Tan Yang Autism Center at Harvard University and the Howard Hughes Medical Institute Fellowship from the Helen Hay Whitney Foundation. X.Q. is supported by the Howard Hughes Medical Institute Fellowship from the Helen Hay Whitney Foundation. I.A. is supported by grant no. T32GM144273 from NIGMS/NIH. R.E.A. is supported by Autism Speaks postdoctoral fellowship 13008. L.A.P. is supported by the grant no. R01HG003988 from NHGRI/NIH. C.A.W. is an Investigator of the Howard Hughes Medical Institute. The content is solely the responsibility of the authors and does not necessarily represent the official views of the National Institute of Health. This study makes use of data generated by the DECIPHER community. A full list of centers who contributed to the generation of the data is available from https://deciphergenomics.org/about/stats and via email from. Funding for the DECIPHER project was provided by Wellcome [grant number WT223718/Z/21/Z]. The clinicians who carried out the original analysis and collection of the data from the DECIPHER community bear no responsibility for the further analysis or interpretation of the data.

## Author Contributions

### Taehwan Shin

Conceptualization, Methodology, Formal analysis, Investigation, Writing - Original Draft, Writing - Review & Editing, Visualization. **Janet H.T. Song**: Conceptualization, Methodology, Formal Analysis, Investigation, Writing-Original Draft, Writing - Review & Editing, Visualization. **Michael Kosicki**: Investigation, Methodology, Writing - Original Draft, Writing - Review & Editing. **Connor Kenny**: Investigation. **Samantha G. Beck**: Investigation. **Lily Kelley**: Investigation. **Xuyu Qian**: Methodology, Investigation. **Julieta Bonacina**: Investigation. **Frances Papandile**: Investigation. **Irene Antony**: Investigation. **Dilenny Gonzalez**: Investigation. **Julia Scotellaro**: Investigation. **Evan Bushinsky**: Investigation. **Rebecca E. Andersen**: Methodology. **Eduardo Maury**: Formal Analysis. **Len A. Pennacchio**: Methodology, Resources, Writing - Review & Editing. **Ryan N. Doan**: Conceptualization, Methodology, Formal analysis, Investigation, Resources, Writing - Original Draft, Writing - Review & Editing. **Christopher A. Walsh**: Conceptualization, Methodology, Resources, Writing - Original Draft, Writing - Review & Editing.

## Materials and Methods

### Cell lines

Neuro2A (N2A) cells (ATCC, cat #CCL-131) were grown in 10% fetal bovine serum and 1% Penicillin-Streptomycin in Dulbecco’s Modified Eagle Medium with L-Glutamine, 4.5g/L Glucose and Sodium Pyruvate (Fisher, cat #MT10013CV). We used a modified version of the male iPSC line WTC11 that contained stably integrated cassettes of a dox-inducible NGN2 and a degron-based inducible dCas9-KRAB (Tian et al., 2019). Both cell lines were maintained in a 5% CO2 incubator at 37°C.

### Mice

Transient transgenic mice were generated using plasmids containing either the human or chimpanzee versions of HAR3091 or HAR3094 located upstream of a minimal promoter driving a *lacZ* reporter gene. These constructs were generated by Vectorbuilder (VB210119-1206gb, VB210119-1208wrc, VB201008-1098whx, VB201020-1677ynn). Pronuclear injections of these constructs were performed in mice by the Mouse Engineering Core at Dana Farber / Harvard Cancer Center or by Cyagen (Santa Clara, CA). Mouse embryos were harvested at E14.5, bisected, and stained for lacZ expression. Embryos were cleared in 30% sucrose-PBS for imaging. Embryos with successful transgene insertions were determined by PCR for the *lacZ* gene. All animal experiments conformed to the guidelines approved by the Children’s Hospital Animal Care and Use Committee.

### Human subjects

From ASD families available through the NIMH repository, we processed 5551 samples (1911 probands) from likely simplex families, and 2277 samples (660 probands) from multiplex families in the Autism Genetic Resource Exchange (AGRE). Variant call format (VCF) files from whole-genome sequencing were obtained for the Homozygosity Mapping Collaborative for Autism (HMCA) from dbGaP phs001894.v1.p1. VCF files for SSC were obtained from SFARI. The number of male and female human subjects in each cohort is indicated in Fig. 4A. Research on human samples was conducted with approval of the Committee on Clinical Investigation at Boston Children’s Hospital.

### Selection of HARs, CNEs, VEs

The set of 3171 HARs examined in this study were selected from a number of different studies that identified HARs separately (Pollard et al., 2006; Bird et al., 2007; Prabhakar et al., 2008; Bush and Lahn, 2008; Lindblad-Toh et al., 2011; Gittelman et al., 2015). Identified HARs that overlap were merged.

VEs were selected from active enhancers from the VISTA Enhancer browser (Visel et al., 2007). The enhancers were filtered for activity in the brain at E11.5, the time point evaluated by the VISTA enhancer group, and because many VEs are very long (greater than 1kb), VEs were subdivided. Only regions that contained at least mildly species-conserved bases (phastCons > 0.57) based on the 100-way vertebrate alignment from the UCSC Genome Browser (Navarro Gonzalez et al., 2021) were selected, with regions 50bp or closer merged to form a single element.

CNEs were selected based on epigenetic datasets, species conservation, and population constraint metrics. CNEs were filtered for conserved genomic regions, defined as having a >400 log-odds of being conserved using phastCons with the Viterbi setting (Siepel et al., 2005). Additionally, CNEs were required to have an enhancer-associated chromatin state (EnhG, Enh, or EnhBiv) based on ChromHMM (Kundaje et al., 2015) in neurospheres, fetal brain, or adult brain. Any elements that were annotated as exonic or splicing were filtered out. Furthermore, no more than 2% of bases in CNEs could have variants in Gnomad (Karczewski et al., 2020).

### Capture-based Massively Parallel Reporter Assay (caMPRA) design, capture, and construction

Molecular inversion probes were designed to capture ∼500bp regions using the MIPgen program (Boyle et al., 2014) to design targeting arms for the probes. Repetitive regions were masked prior to targeting. Flanking sequence was used to introduce AsiSI, PspXI, and SfiI restriction enzyme sites (NEB, cat #R0630L, R0656L, R0123L), along with a 10bp barcode for each probe. As targeted regions vary in length and many elements are longer than 500bp, probes were designed to double tile the bases of each element. MIPs were synthesized by Customarray, Inc (Redmond, WA).

The synthesized MIP oligos were amplified, amplification arms cleaved using MlyI (NEB, cat #R0610L), and purified using Ampure XP beads (Beckman Coulter) at 1.8x volume of the reaction mix. 15ng of the amplified MIP probes were then hybridized to 500ng of DNA from sample NA12878 (Coriell Institute) for 24 hours in 10x Ampligase buffer. The sequences in between the MIP targeting arms were captured by synthesis using Phusion DNA Polymerase (Thermo Fisher, cat #F530L) and circularized using Ampligase (VWR, cat #A3210K) at 60°C for 1 hour. Template DNA and uncaptured DNA were degraded using Exonuclease I (Thermo Fisher, cat #EN0582) and Exonuclease III (Thermo Fisher, cat #EN0191) at 37°C for 40 minutes and inactivated at 95°C for 5 minutes. The captured circles were then amplified using Phusion DNA Polymerase High Fidelity Master Mix (Thermo Fisher, cat #F531L), using primers that added SfiI restriction sites. Amplified, captured sequences were purified using Ampure XP beads at 0.65X to size select and remove unwanted shorter fragments.

The captured sequences and the pMPRA1 construct (Melnikov et al., 2014) (Addgene, cat #49349) were digested with SfiI and ligated using T4 DNA Ligase (Thermo Fisher, cat #EL0014) at 16°C overnight. The ligated construct was purified and concentrated using the QIAquick Nucleotide Removal Kit (Qiagen, cat #28306). The ligated constructs were either transformed into 60 vials of Top10 chemically competent cells (Thermo Fisher, cat #C404006) and cultured overnight at 37°C in 200ml of LB/Ampicillin, or transformed into 1 vial of MegaX DH10B T1R Electrocompetent Cells (Thermo Fisher, cat #C640003) and plated on LB/Ampicillin agar plates (Molecular Devices X6023 BIOASSAY TRAYS; Fisher Scientific, cat #NC9372402) overnight at 37°C. Plasmid DNA was extracted the following day using the Qiagen Plasmid Maxi Kit (Qiagen, cat #12162). This plasmid library containing the captured sequences and a modified pMPRAdonor2 (Melnikov et al., 2014) (Addgene, cat #49353) containing an AsiSI site were then digested using AsiSI and PspXI, and the fragment containing the minimal promoter and luciferase gene from pMPRAdonor2 was cloned into the plasmid library containing the captured sequences between the captured element and the barcode. This final construct was then transformed, cultured, and harvested as above.

For the random mutagenesis of HARs, a 25bp barcode was used. We performed error-prone PCR using the GeneMorph II Random Mutagenesis Kit (Agilent, cat #200550) on the amplified, captured sequences. Based on the error rate of the Mutazyme II polymerase and PCR yield, we performed 7 cycles of error-prone PCR with 20ng of input captured sequence, with 25bp random barcode reverse primer. These mutagenized sequences were then cloned into the modified pMPRA1 construct as described above. In order to associate the mutagenized sequence with the random barcode, the cloned plasmid library containing the captured sequences was PCR amplified with primers containing sequencing adapters and sent out for 2x250bp sequencing on HiSeq Instruments at Psomagen.

### Cell culture and transfection for caMPRA

N2A cells (ATCC, cat #CCL-131) were cultured in DMEM with L-Glutamine, 4.5g/L Glucose and Sodium Pyruvate (Fisher, cat #MT10013CV) with 10% fetal bovine serum and 1% penicillin/streptavidin at 37°C. Cells were maintained in 10cm TC-treated plates and split 1:5 every 4 days or when confluent with 0.25% (w/v) Trypsin – 0.53mM EDTA (Fisher, cat #25200-114). To minimize confounding due to passage number, we limited passage numbers to P3-6. For transfections, N2a cells were transfected at 70% confluency using Lipofectamine LTX with PLUS reagent (Thermo Fisher, cat #15338100) with 15ug of caMPRA plasmid, and cells were incubated with the transfection mix for 24 hours. After 1 day, media was changed. Cells were harvested either 1 day or 3 days after transfection. Cell pellets were washed with 1x PBS, and mRNA was extracted using the Dynabead mRNA Direct kit (Thermo Fisher, cat #61012), according to the manufacturer’s instructions. mRNA was reverse transcribed using Superscript VILO Master Mix with EZ DNase (Thermo Fisher, cat #11766050). caMPRA barcodes were extracted using PCR amplification with primers containing illumina adapters for both the cDNA and plasmid pools and sent out for 150bp sequencing using Hiseq instruments at Psomagen (Rockville, MD).

### Targeted sequencing of NIMH cohort

From the ASD families available through the NIMH repository, we processed 5551 samples (1911 probands) from likely simplex families, and 2277 samples (660 probands) from multiplex families in the Autism Genetic Resource Exchange (AGRE). The likely simplex families include families with one affected proband and no siblings and families with one affected proband and one or more unaffected siblings. Molecular inversion probes (MIPs) were designed, synthesized, and amplified as described above with the following changes: MIPs were designed to capture ∼240 bp regions and were hybridized to a pool of DNA from NIMH samples. The purified library was quantified using a tapestation and sequenced at 2x150bp by Psomagen (Rockville, MD).

### LacZ enhancer reporter assay in transgenic mice with random integration sites

We cloned either the human or chimpanzee versions of HAR3091 or HAR3094 located upstream of a minimal promoter driving a *lacZ* reporter gene with Vector-builder (VB210119-1206gb, VB210119-1208wrc, VB201008-1098whx, VB201020-1677ynn). Pronuclear injections of these constructs were performed in mice by the Mouse Engineering Core at Dana Farber / Harvard Cancer Center or by Cyagen (Santa Clara, CA). Mouse embryos were harvested at E14.5, bisected, and stained for lacZ expression. Embryos were cleared in 30% sucrose-PBS for imaging. Embryos with successful transgene insertions were determined by PCR for the lacZ gene. Because these mice are analyzed at F0, lacZ expression is dependent on the distribution and number of cells that integrate the reporter construct and the genomic location of the integration. Consequently, we expect that expression patterns driven by the sequence of interest rather than by the integration site will be consistently observed in multiple embryos and examined at least 10 PCR-positive embryos per construct to account for this variability.

### LacZ enhancer reporter assay in transgenic mice with site-specific integration

Transgenic E11.5 mouse embryos were generated as described previously (Osterwalder et al., 2022). Briefly, superovulating female FVB mice were mated with FVB males and fertilized embryos were collected from the oviducts. Regulatory elements sequences were synthesized by Twist Biosciences. Inserts generated in this way were cloned into the donor plasmid containing minimal Shh promoter, lacZ reporter gene and H11 locus homology arms (Addgene, cat #139098) using NEBuilder HiFi DNA Assembly Mix (NEB, cat #E2621). The sequence identity of donor plasmids was verified using long-read sequencing (Primordium). Plasmids are available upon request. A mixture of Cas9 protein (Alt-R SpCas9 Nuclease V3, IDT, cat #1081058, final concentration 20 ng/μL), hybridized sgRNA against H11 locus (Alt-R CRISPR-Cas9 tracrRNA, IDT, cat #1072532 and Alt-R CRISPR-Cas9 locus targeting crRNA, gctgatggaacaggtaacaa, total final concentration 50 ng/μL) and donor plasmid (12.5 ng/μL) was injected into the pronucleus of donor FVB embryos. The efficiency of targeting and the gRNA selection process is described in detail in (Osterwalder et al., 2022). Embryos were cultured in M16 with amino acids at 37°C, 5% CO_2_ for 2 hours and implanted into pseudopregnant CD-1 mice. Embryos were collected at E11.5 for lacZ staining as described previously (Osterwalder et al., 2022). Briefly, embryos were dissected from the uterine horns, washed in cold PBS, fixed in 4% PFA for 30 min and washed three times in embryo wash buffer (2mM MgCl2, 0.02% NP-40, and 0.01% deoxycholate in PBS at pH 7.3). They were subsequently stained overnight at room temperature in X-gal stain (4mM potassium ferricyanide, 4mM potassium ferrocyanide, 1mg/mL X-gal and 20mM Tris pH 7.5 in embryo wash buffer). PCR using genomic DNA extracted from embryonic sacs digested with DirectPCR Lysis Reagent (Viagen, cat #301-C) containing Proteinase K (final concentration 6 U/mL) was used to confirm integration at the H11 locus and test for presence of tandem insertions (see (Osterwalder et al., 2022) for details). Only embryos with donor plasmid insertion at H11 were used. The stained transgenic embryos were washed three times in PBS and imaged from both sides using a Leica MZ16 microscope and Leica DFC420 digital camera.

### CRISPR inhibition in iPSC-derived neurons

Guide RNAs (gRNAs) were designed to target the middle 200bp interval of HAR3091 and HAR3094 using GuideScan (Perez et al., 2017) with a specificity score cut-off > 0.2. Non-targeting control (NTC) gRNAs and gRNAs targeting the *IL1RAPL1* transcription start site (TSS) are from (Horlbeck et al., 2016). gRNA sequences can be found in Table S5. gRNAs were cloned into pBA904 (RRID: Addgene 122238), as previously described (Replogle et al., 2020). Lentivirus was made for each gRNA by ultracentrifugation.

We used a modified version of the iPSC line WTC11 that contained stably integrated cassettes of a dox-inducible NGN2 and a degron-based inducible dCas9-KRAB (Tian et al., 2019). This modified WTC11 line was differentiated into neurons by inducing NGN2 expression as previously described (Chen et al., 2020). Twice the suggested number of cells (e.g. 2*x*10^5^ cells per well in a 24-well plate) were plated at D0 to account for incomplete lentiviral infection, and immediately after plating, lentivirus was added at MOI 0.7 (so that ∼50% of cells would be infected with lentivirus). dCas9-KRAB expression was induced by the addition of 20μM trimethoprim (Sigma Aldrich, cat #92131) from D0 until the neurons were collected at D18, and 1 μg/ml puromycin was added from D2-D7 to select for infected neurons. RNA was extracted at D18 using the Directzol RNA Microprep Kit (Zymo, cat #R2062) per the manufacturer’s instructions, and cDNA was synthesized from RNA with the SuperScript VILO cDNA Synthesis Kit (Thermo Fisher, cat #11756050). RT-qPCR was performed using Brilliant II SYBR Green Low ROX qPCR Master Mix (Agilent, cat #600830) on a CFX384 Touch Real-Time PCR Detection System (BioRad) for IL1RAPL1 and GAPDH (primer sequences in Table S5) in triplicate.

### Luciferase assays

The wild-type (WT) and mutant sequences of for HAR 3091 and 3094 were generated through PCR amplification from Promega Control Male human DNA (Promega, cat #G1471) and proband genomic DNA using primers containing unique restriction sites for directional cloning into a minimal promoter pGL4.25 luciferase. Two families harboring the same variant in HAR3091 (chrX:29275879, G>T) were amplified independently and cloned into separate plasmids containing the same variant. Plasmids were transformed into Top10 chemically competent cells (Thermo Fisher, cat #C404006). Genotypes and structures of the final plasmids were confirmed using Sanger sequencing. Plasmids (75ng) were co-transfected along with control Renilla (25ng) into mouse neuroblastoma Neuro-2a cells (N2a) (ATCC, cat #CCL-131) in 96-well plates using Lipofectamine 3000 (Thermo Fisher, cat #L3000015). Luciferase activities were assessed 48 hours post-transfection using standard procedures for the dual-luciferase reporter assay system (Promega, cat #E1960). The firefly luciferase activity was normalized using Renilla activity for each well of the 96-well plates. The luciferase activity measurements were performed with 8 replicates. Data from the two plasmids generated from different families with the same variant in HAR3091 yielded similar results, and are shown as one variant (Fig. 5D, Fig. S15).

### Assessing accessibility and epigenetic marks in human tissue

ChromHMM annotations from the Epigenomics Roadmap Project (Kundaje et al., 2015) were overlapped with HARs, CNEs, and VEs using bedtools intersect (Quinlan and Hall, 2010) to identify the number of elements in each class that were annotated as active (TssA, TssAFlnk, TxFlnk, Tx, TxWk, EnhG, Enh, TssBiv, BivFlnk, or EnhBiv in the 15-state model) in each assessed tissue. For cell type-specific annotations in the adult brain, scTHS-seq data was used (Lake et al., 2018) to determine accessibility in different brain cell types for HARs, VEs, and CNEs.

### TF binding analysis

We examined HARs, VEs, and CNEs, as well as matched background sequences generated using BiasAway (Khan et al., 2021), for transcription factor binding sites. PWMSCAN (Ambrosini et al., 2018) was used to scan sequences for motifs from the JASPAR motif database (Castro-Mondragon et al., 2022) to identify potential transcription factor binding sites. To control for false positives, a p-value cutoff of 10^−4^ was used. The presence of motifs was then aggregated and the enrichment of specific motifs in HARs, VEs, or CNEs compared to matched background sequences was determined. P-values for enrichment were generated using the hypergeometric test and were adjusted for multiple hypothesis testing with the Benjamini-Hochberg correction. Gene set enrichment analysis for TFs with motifs in HARs, VEs, and CNEs was performed using clusterProfiler (Yu et al., 2012) and adjusted for multiple hypothesis testing with the Benjamini-Hochberg correction.

In addition to motif-based matching, HARs, VEs, and CNEs were also annotated using DeepSEA (Zhou et al., 2018) using the online DeepSEA server. We used the Beluga model that was trained on 2,002 chromatin features. TF ChIP-seq features were selected for, and only those with an e-value (defined as the expected proportion of common SNPs with a larger predicted effect) of less than 0.05 were interpreted as associated with the element.

### Analysis of genes near HARs, VEs, and CNEs

Gene ontology analysis was performed with GREAT (McLean et al., 2010) with the binomial test at 5% FDR. The binomial test at 5% FDR were also used to assess whether HARs, VEs, and CNEs are enriched near disease-associated genes. HARs, VEs, and CNEs were assigned to nearby genes as previously described (McLean et al., 2010). We separated genes implicated in severe, developmental disorders from the DECIPHER consortium (v. 13 7 2022) (Firth et al., 2009) based on the phenotypes of the affected individuals. If affected individuals had phenotypes in multiple body systems, affected genes were assigned to all affected body systems. We also examined autism-associated genes from the SFARI database (Abrahams et al., 2013).

To examine whether autism-associated genes and genes near HARs, VEs, or CNEs were enriched for dosage-sensitive genes, we examined pLI and LOEUF scores (Lek et al., 2016; Karczewski et al., 2020). pLI > 0.9 and low LOEUF scores indicate loss-of-function intolerance. The hypergeometric test was used to test whether ASD-associated genes from the SFARI database and genes near HARs, VEs, or CNEs were enriched for genes with pLI > 0.9 at 5% FDR. The Wilcoxon rank-sum test was used to test whether the LOEUF scores of ASD-associated genes or genes near HARs, VEs, or CNEs differed from LOEUF scores for all genes at 5% FDR.

### caMPRA analysis

To count barcodes to assess regulatory activity, sequencing data from the plasmid DNA and cDNA libraries described above were processed with cutadapt to remove adapters (Martin, 2011). Barcodes were extracted using UMI-tools (Smith et al., 2017) and reads were mapped using bwa mem (Li and Durbin, 2009). Reads were assigned to caMPRA probes using featureCounts (Liao et al., 2014). The 10bp barcode were clustered to recover barcodes with small sequencing errors using the multiplexed version of the UMI-tools directional method.

Barcodes for plasmid and cDNA samples were normalized to a barcode per million format to remove bias due to sequencing coverage. Each cDNA barcode was normalized to the barcode count in the plasmid pool, and *log*_2_ transformed. Only barcodes that were found in the plasmid pool and all 5 cDNA replicate pools were used in the analysis. After filtering, we were able to examine 2932 HARs, 1702 VEs, and 5155 CNEs. Elements were considered active if at least one probe overlapping that element was active in the assay (Fig. 2B, Fig. S6B). The proportion of active probes (sequences) is shown in Fig. S6A, and full results are detailed in Table S2.

### Analysis of caMPRA data from random mutagenesis

Analysis of the plasmid and cDNA barcode pools was performed as described above. Variants from each caMPRA probe were called using bcftools mpileup (Li, 2011), and associated with the appropriate barcode. Sequences captured using MIPs may include regions that flank the sequence of interest. Mutagenized sequences that only included variants in the flanking sequence were excluded. Variants found in NA12878 compared to the reference genome were filtered out. The correlation between replicate experiments (Fig. S8) was assessed prior to removing mutagenized sequences that only included variants in the flanking sequence.

### MIP sequencing pre-processing and variant calling

Analysis of targeted sequencing was performed using a custom pre-processing pipeline combined with GATK-based variant calling. Briefly, sequenced reads were trimmed for adapters using cutadapt (Martin, 2011). UMI-tools (Smith et al., 2017) was used to extract the 5bp unique molecular index (UMI), and reads were mapped to the human genome (hg19) using bwa mem (Li and Durbin, 2009). Reads that mapped off-target compared to the intended target were filtered out. The extension and ligation arms (the targeting arms) were clipped off the mapped reads using bamclipper (Au et al., 2017). Samtools was used to remove multimapping reads, unpaired/broken read pairs, and unmapped reads (Li et al., 2009). UMI-tools was used to collapse sequences based on UMI sequence. Finally, sequences were base recalibrated using GATK base recalibrator, and variants were joint-called using the GATK Haplotype caller and suggested GATK best practices (Van der Auwera et al., 2013). After targeted sequencing and processing, we were able to resolve HARs in 6464 individuals, VEs in 5273 individuals, and CNEs in 5983 individuals.

### Variant filtering, classification, and analysis

The HMCA consanguineous cohort was filtered for variants of AD > 2, DP > 10, GQ > 20. For the targeted sequencing of the NIMH cohort, variants were required to have a minimum of 10x coverage and GQ > 20 and AD > 4. Only variants produced from collapsed reads were used for accuracy. The Simon Simplex Collection (SSC) cohort of 8,186 individuals was filtered to remove alleles with AD < 3, DP < 5, and GQ < 20.

For recessive variant analysis, our definition included homozygous, compound heterozygous, and hemizygous variants (specifically in male individuals for the X chromosome). Because hemizygous variants are much more likely to appear, we performed our analysis with each sex separately when examining genome-wide rates.

In order to enrich for functionality, we created a classification that uses an ensemble of different conservation-based variant effect predictors – GERP++ (Davydov et al., 2010), CADD (Rentzsch et al., 2019), DANN (Quang et al., 2015), FATH-MMnc (Shihab et al., 2015) -to annotate variants and base positions. Variants were filtered to exclude those within exonic regions of protein-coding genes (based on RefSeq and Gencode v28). For variants that fall within either the UTRs, within 1 kb upstream of a transcriptional start site, or within a predicted promoter element from the Eukaryotic Promoter Database (Dreos et al., 2015), these variants must overlap a conserved element from the 100-way phastCons from the UCSC genome browser. All variants were filtered for GERP > 2 and (CADD > 15 or DANN > 0.85 or FATHMMnc > 0.85).

The variant contributions of rare germline events were assessed for rare, recessive and *de novo* predicted damaging variants identified in individuals with ASD and healthy familial controls. Statistical testing of variant contributions was performed as follows. First, the odds ratio (*OR*), standard error, and 95% confidence intervals were calculated using the approach described by (Altman, 1991): 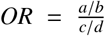, where a = number of cases with variants, b = number of cases without variants, c = number of controls with variants, and d = number of controls without variants. The standard errors of the log odds ratio were calculated using the following formula: 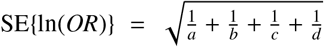. The 95% confidence intervals were determined using 95% CI = exp(ln(*OR*) + 1.96× SE ln(*OR*)). P-values of the ORs were calculated under the assumption of the deviation from a normal distribution, using: 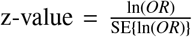. The allele frequency (AF) cut-off for statistical analysis was set at either *AF* < 0.005 (HMCA and SSC) or *AF* < 0.001 (NIMH), using the lowest AF where there were >5 predicted damaging variants for cases and controls for each sex. The contribution of variants to ASD risk was estimated as the difference between the rate of rare, recessive variants in cases compared to controls as previously described (Doan et al., 2019).

In cohorts with known elevations of homozygosity that could impact the recessive contribution (e.g., HMCA), we determined the rates of likely benign events at non-conserved sites within gene promoters that have no predicted functional impact under the assumption that these rates should be equivalent in cases and controls due to the lack of selection bias on the sites. Next, the rates of predicted damaging events in cases were reduced proportionally to the excess detected in the non-conserved sites, as was done previously using synonymous rates for recessive protein coding variation (Doan et al., 2019). Following correction for elevated consanguinity, variation contributions and significance were determined, using the above described approach.

### Protein-protein interaction networks

Variants found in HARs, VEs, or CNEs in HMCA and variants found in HARs or VEs in NIMH were aggregated to the level of individual HARs, VEs, and CNEs. HARs, VEs, and CNEs with a numerical excess of variants found in patients compared to controls were associated with nearby genes using GREAT (McLean et al., 2010). A protein-protein interaction network for these nearby genes was constructed using STRING version 11.5 using the online interface with default parameters (Szklarczyk et al., 2019).

### Analysis of CRISPR inhibition in iPSC-derived neurons and luciferase assays

To analyze the CRISPR inhibition data, the quantity of *IL1RAPL1* for each sample was calculated by comparing the Ct value for *IL1RAPL1* to a standard curve of pooled samples and then normalized to the expression of the housekeeping gene *GAPDH* in the same sample. This normalized value was then divided by the normalized quantity of *IL1RAPL1* in samples infected with NTC gRNAs. Each point represented in Fig. 5 and Fig. S14 is from a separate well; each condition was tested in at least 3 different differentiations. The Wilcoxon rank-sum test was used to compare each individual gRNA to the NTC gRNAs, and p-values for gRNAs targeting the same region were combined with Fisher’s method. P-values were adjusted with the Benjamini-Hochberg correction and considered significant at 5% FDR.

For the luciferase assays, the Wilcoxon rank-sum test was used to compare each test sequence to the control sequence, and p-values for each replicate were combined with Fisher’s method. P-values were adjusted with the Benjamini-Hochberg correction and considered significant at 5% FDR.

