## Supplement for "Rare variation in noncoding regions with evolutionary signatures contributes to autism spectrum disorder risk"

**Supplemental Figures**

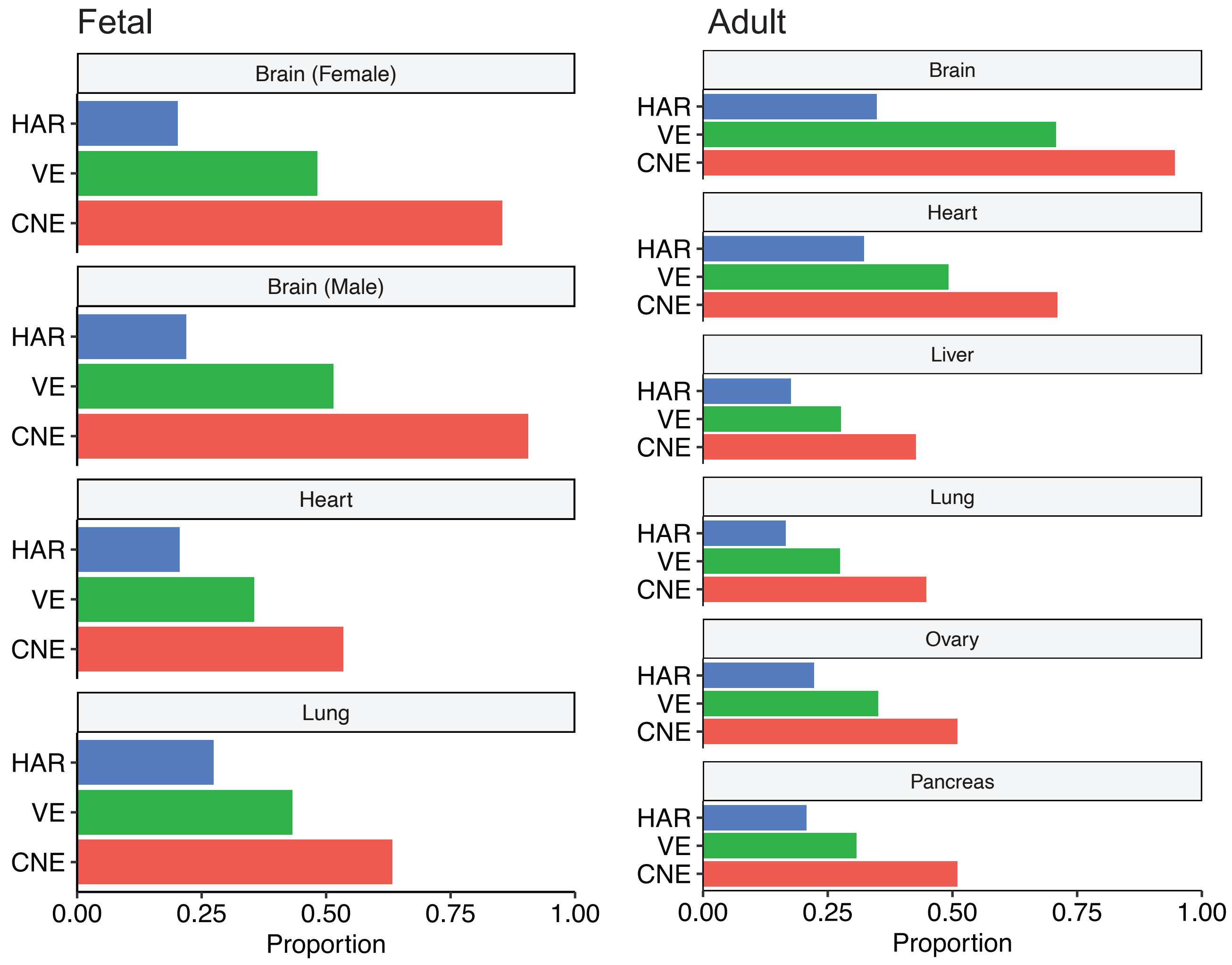


Figure S1: **Proportion of HARs, VEs, and CNEs predicted to be active in fetal (left) and adult (right) tissue by ChromHMM from the Roadmap Epigenomics Project (Kundaje et al., 2015) (Materials and Methods).**

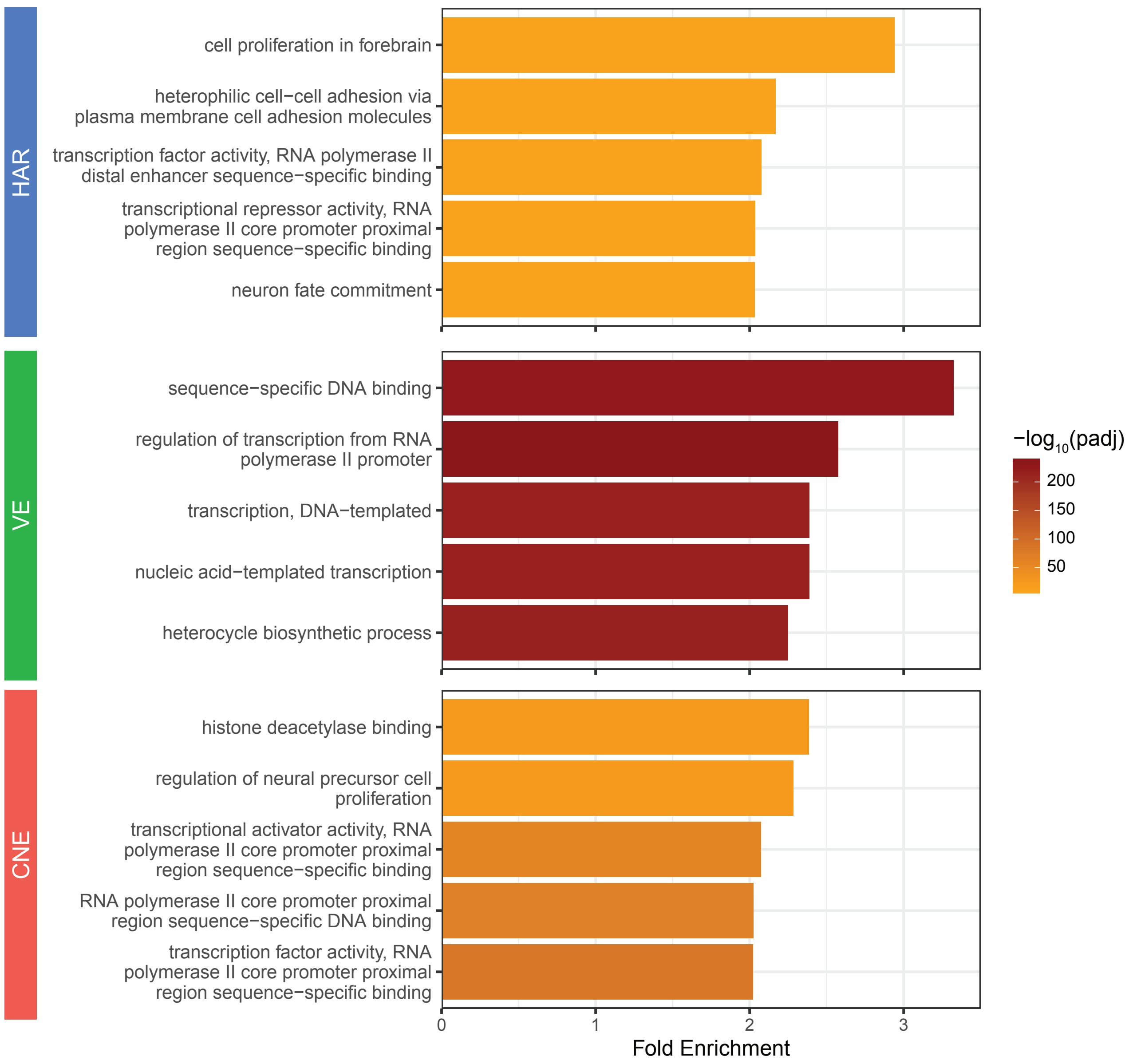


Figure S2: **Gene ontology enrichments for genes near HARs, VEs, and CNEs using GREAT (McLean et al., 2010).** The top five enriched terms from the binomial test are plotted.

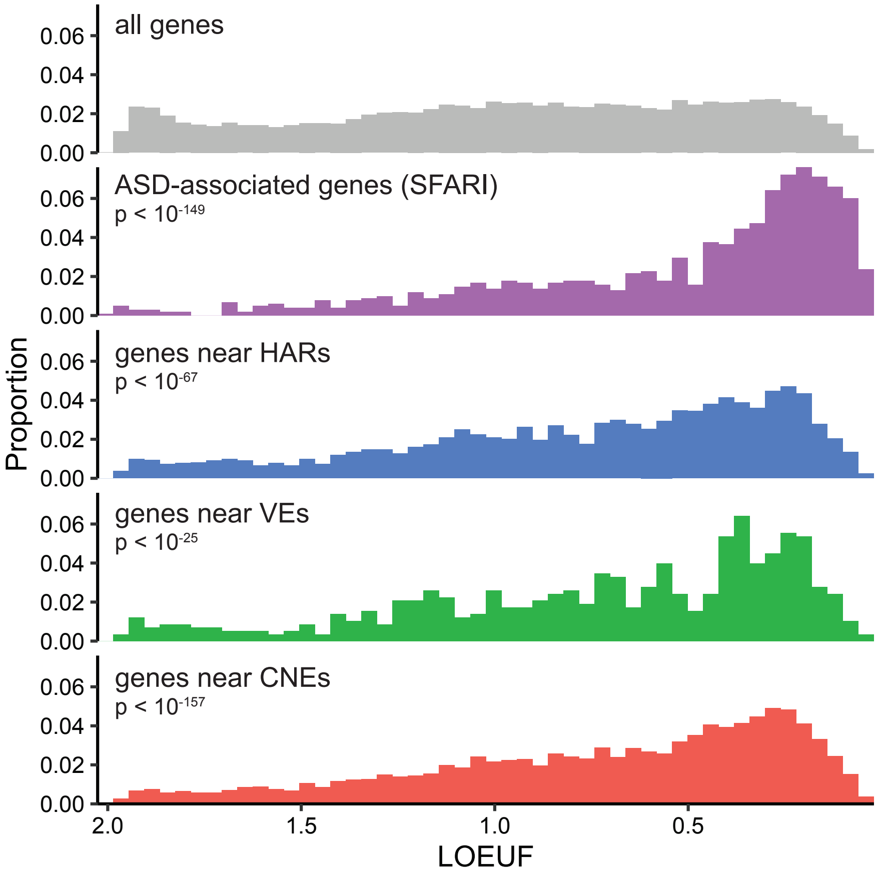


Figure S3: **ASD-associated genes from the SFARI database (Abrahams et al., 2013) and genes near HARs, VEs, and CNEs are enriched for low LOEUF scores compared to all genes.** Genes that are loss-of-function intolerant have low LOEUF scores (Karczewski et al., 2020).

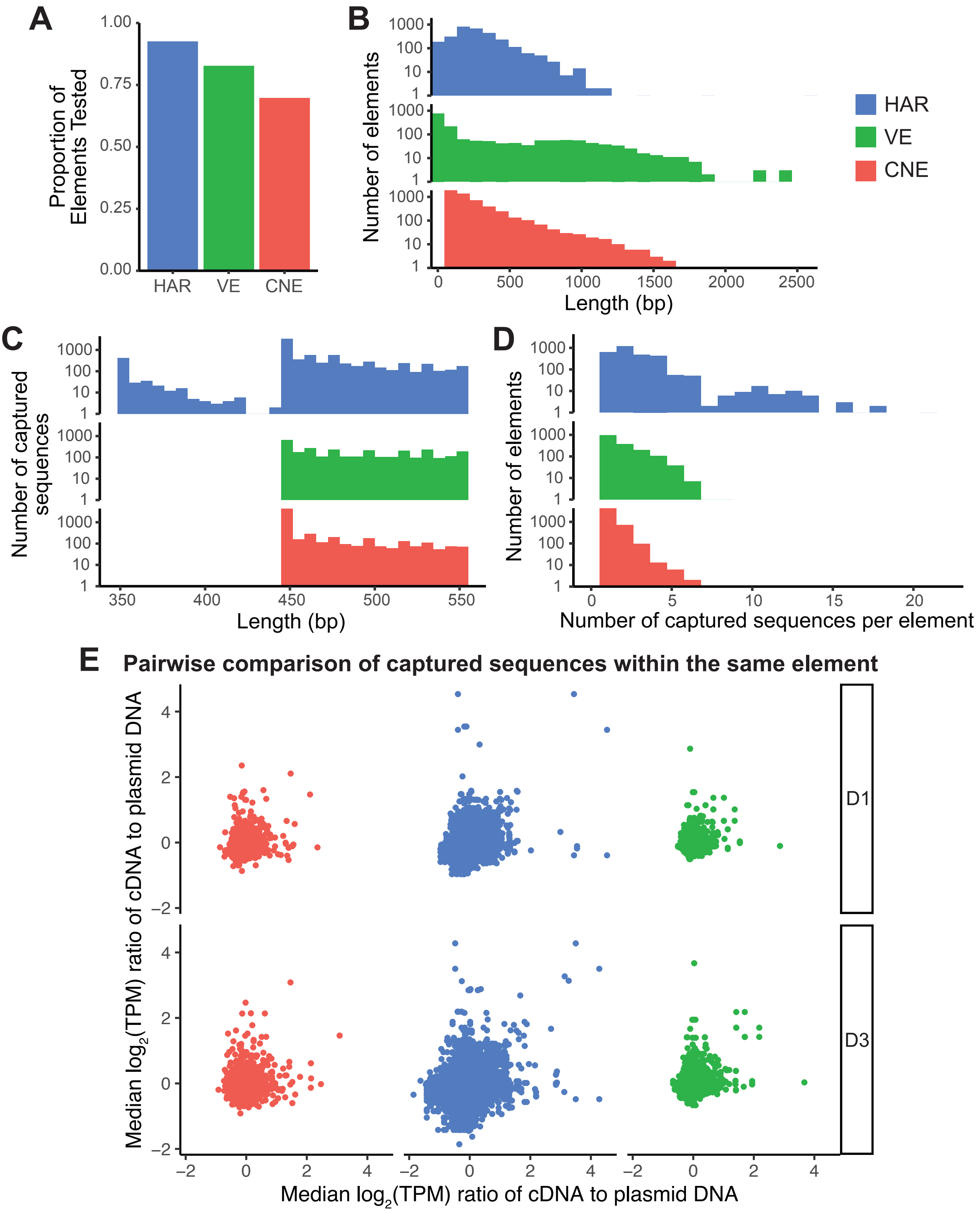


Figure S4: **Features of caMPRA for HARs, VEs, and CNEs.** (A) Proportion of HARs, VEs, and CNEs tested in caMPRA. (B) Distribution of the length of HARs, VEs, and CNEs. (C) Length and number of sequences captured for HARs, VEs, and CNEs. (D) Distribution of the number of captured sequences per HAR, VE, and CNE. (E) Normalized ratio of cDNA to plasmid DNA (enhancer activity) for sequences captured from the same HAR, VE, or CNE. The enhancer activity of captured sequences from the same element is not correlated.

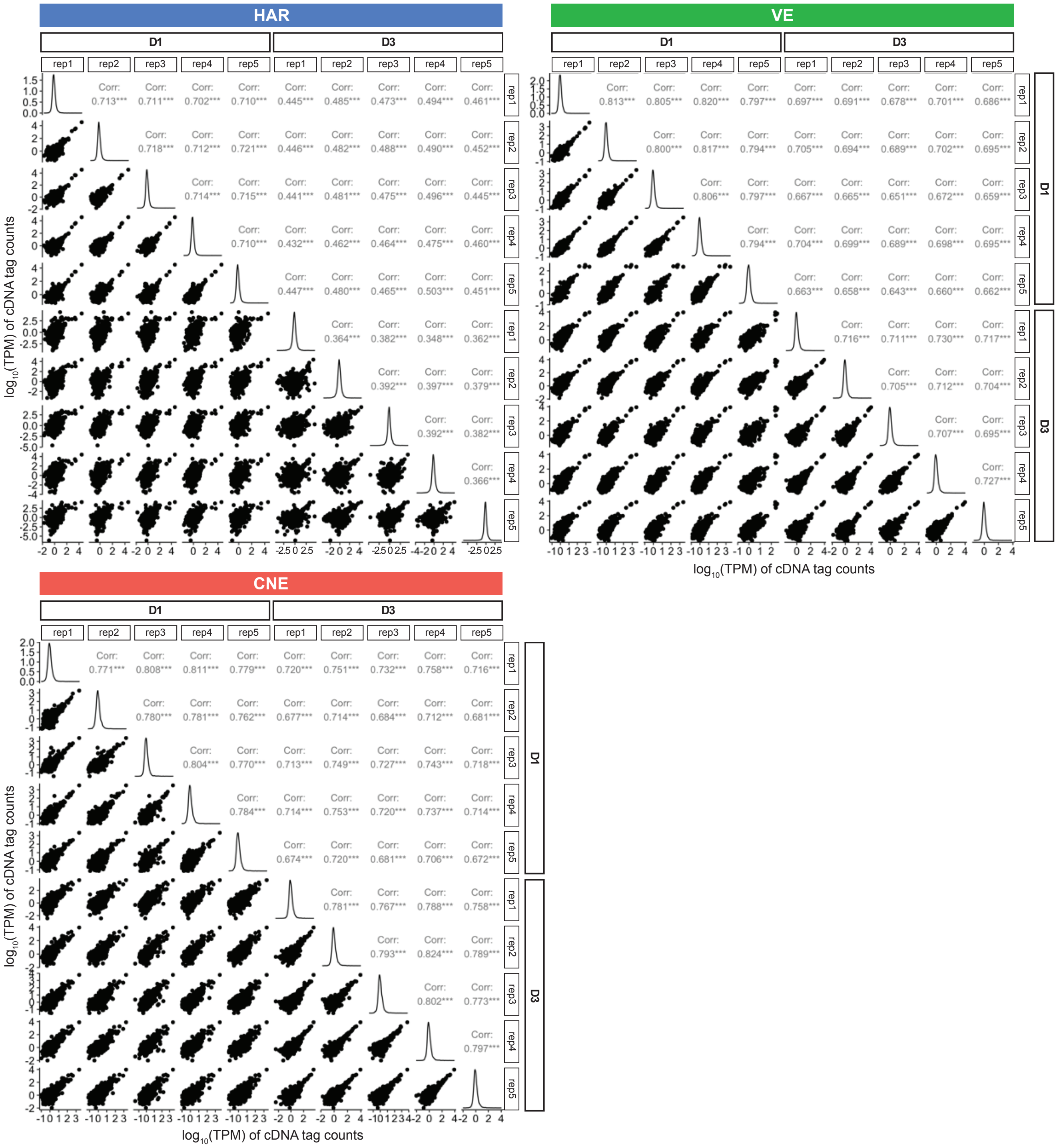


Figure S5: **Enhancer activity in caMPRA of HARs, VEs, and CNEs is well-correlated between replicates and between collection timepoints.**

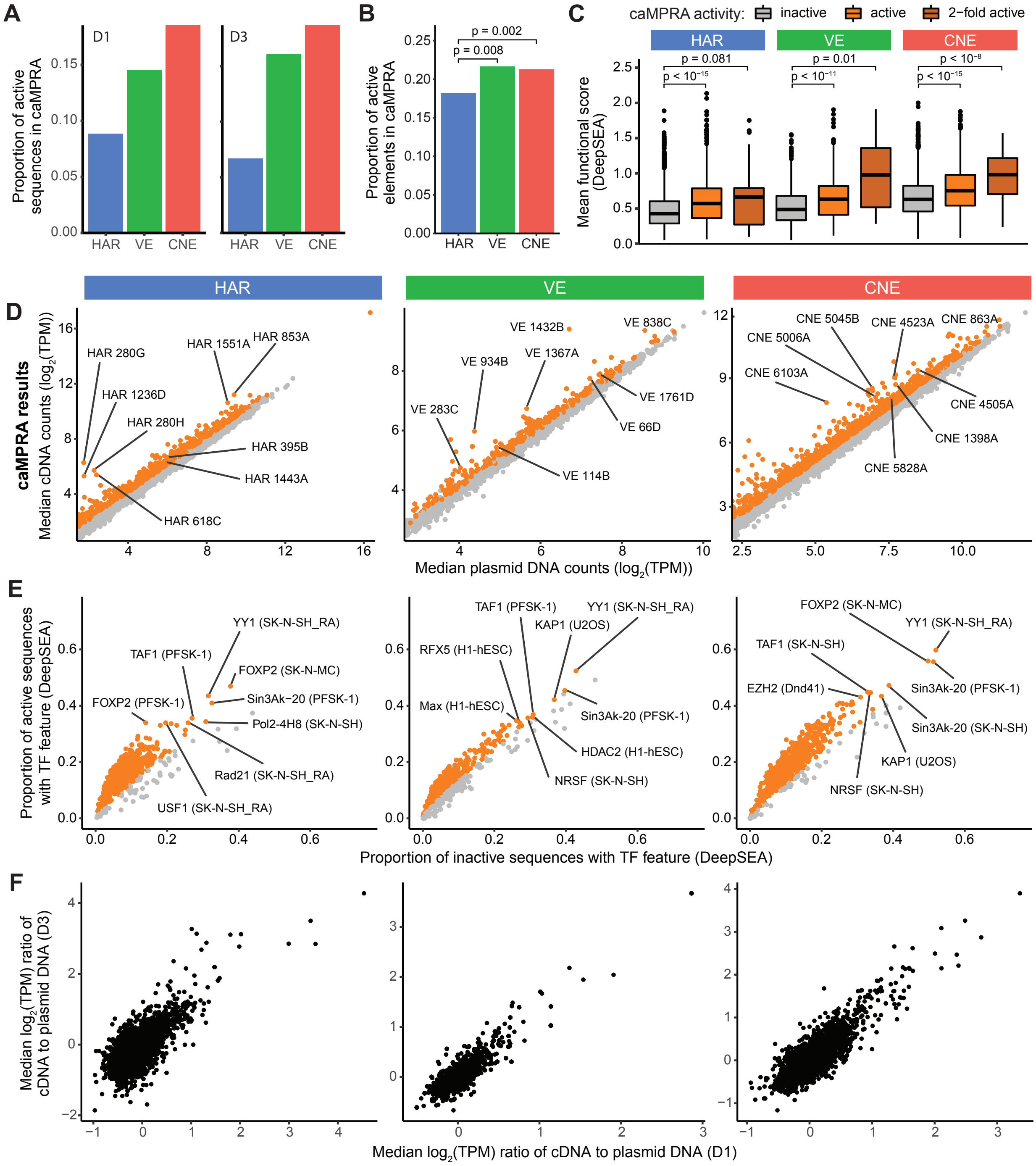


Figure S6: **HARs, VEs, and CNEs display enhancer activity in a capture-based Massively Parallel Reporter Assay (caMPRA).** (A) Proportion of captured sequences that are active in caMPRA for HARs, VEs, and CNEs when assessed one (D1) or three (D3) days after transfection. (B) Proportion of HARs that have enhancer activity in at least one captured sequence in the D1 caMPRA is significantly lower than VEs or CNEs by the chi-square test after FDR correction. (C) Sequences captured from HARs, VEs, and CNEs are classified as inactive, active, or 2-fold active from the D1 caMPRA experiment and compared to their mean functional score from DeepSEA (average of $-log_{10}(e - value)$ for every feature) (Zhou and Troyanskaya, 2015). The level of activity in caMPRA is correlated with the predicted mean functional score from DeepSEA. P-values were determined with the hypergeometric test and adjusted by FDR correction. (D) Normalized cDNA counts vs normalized plasmid counts for sequences captured from HARs, VEs, and CNEs from the D1 caMPRA experiment. Sequences with significant enhancer activity are in orange. (E) TF features were predicted by DeepSEA for each captured sequence. TF features significantly enriched in active sequences in the D1 caMPRA experiment are shown in orange. Representative TF features are marked in the format: TF (cell type). (F) The enhancer activity of captured sequences between the D1 and D3 caMPRA experiments is highly correlated.

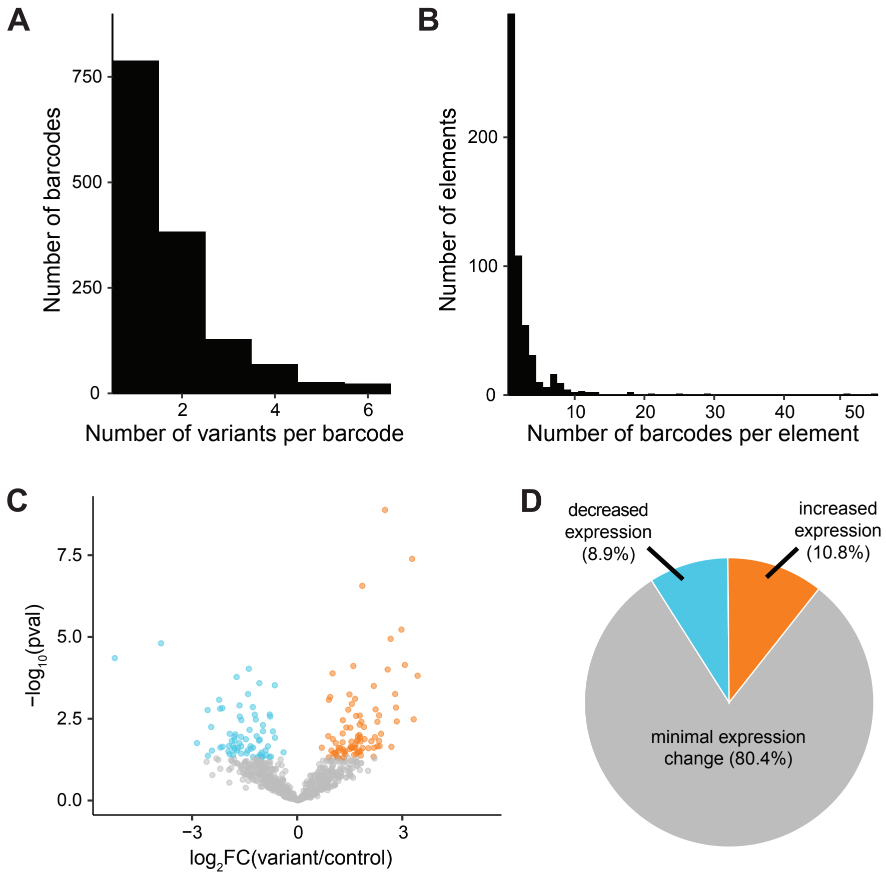


Figure S7: **Single nucleotide variants can modulate enhancer activity.** (A) Number of variants per mutagenized sequence (barcode). (B) Number of barcodes tested per probe. There are at least two designed probes (elements) per HAR. (C) Volcano plot of fold change in expression and adjusted p-value for barcodes that contain only one introduced variant ($n$ = 789). (D) Pie chart of percent of barcodes with decreased expression, increased expression, or no statistically significant change in expression for barcodes that contain only one introduced variant.

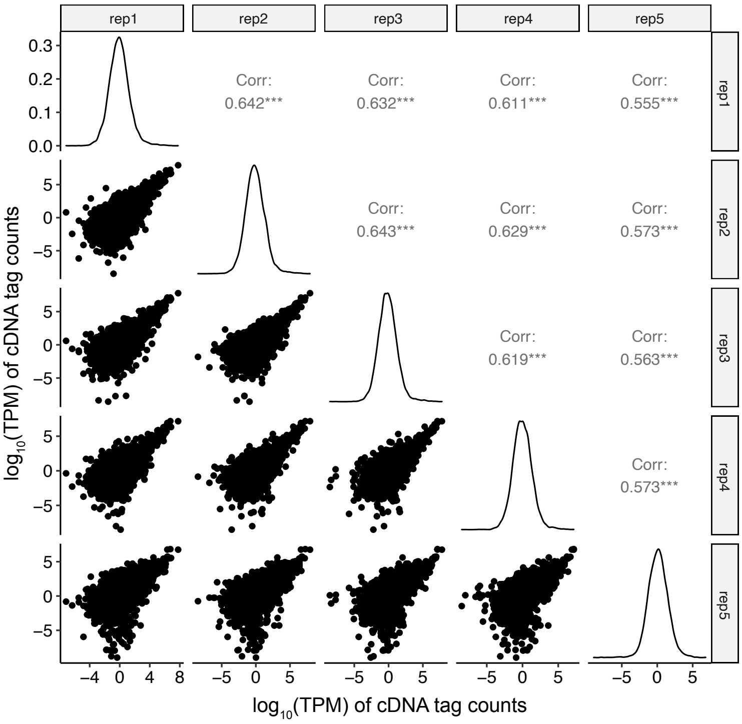


Figure S8: **Enhancer activity in caMPRA of random mutagenesis of HARs is well-correlated between replicates.**

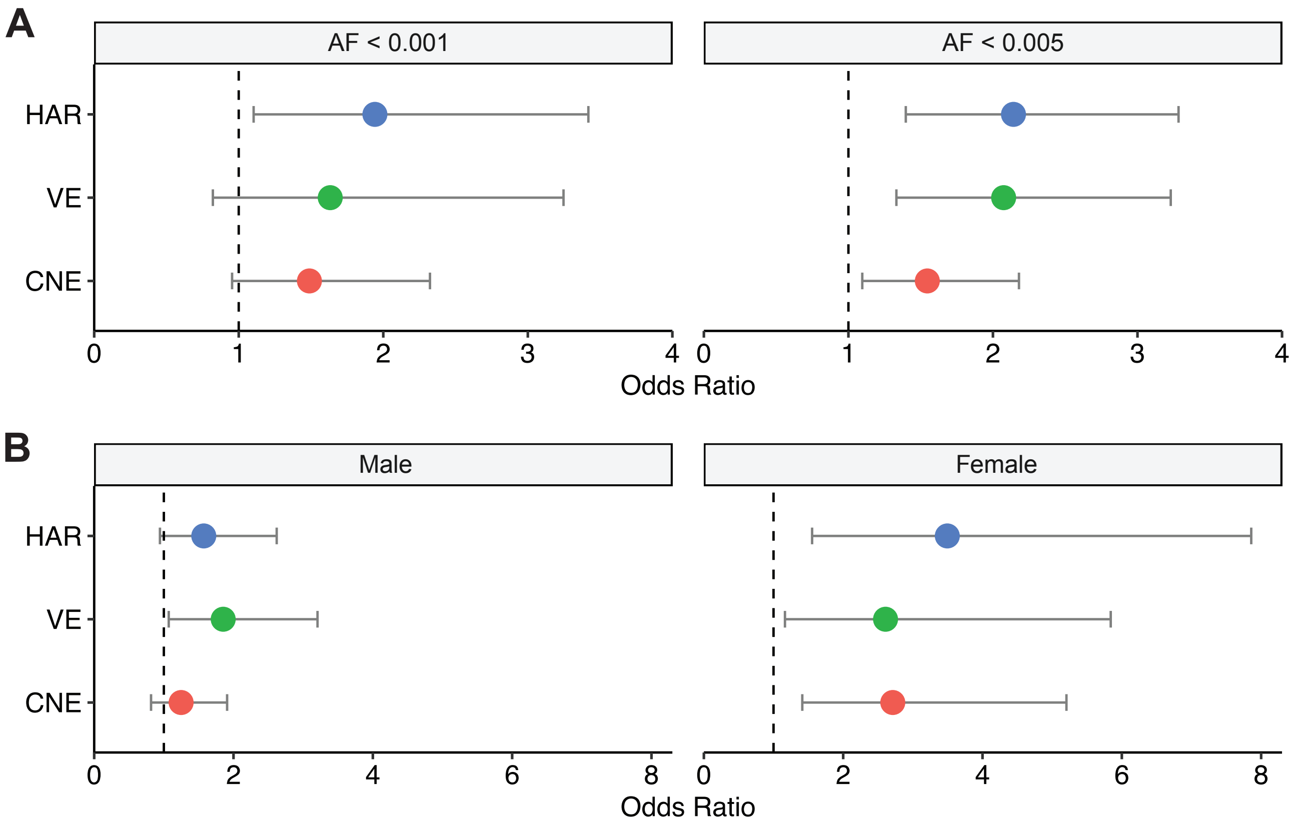


Figure S9: **Odds ratios for rare, recessive variants at conserved bases in cases versus controls in HMCA cohort are consistent across allele frequencies (A) and sexes (B).** (B) is assessed at allele frequency (AF) < 0.005.

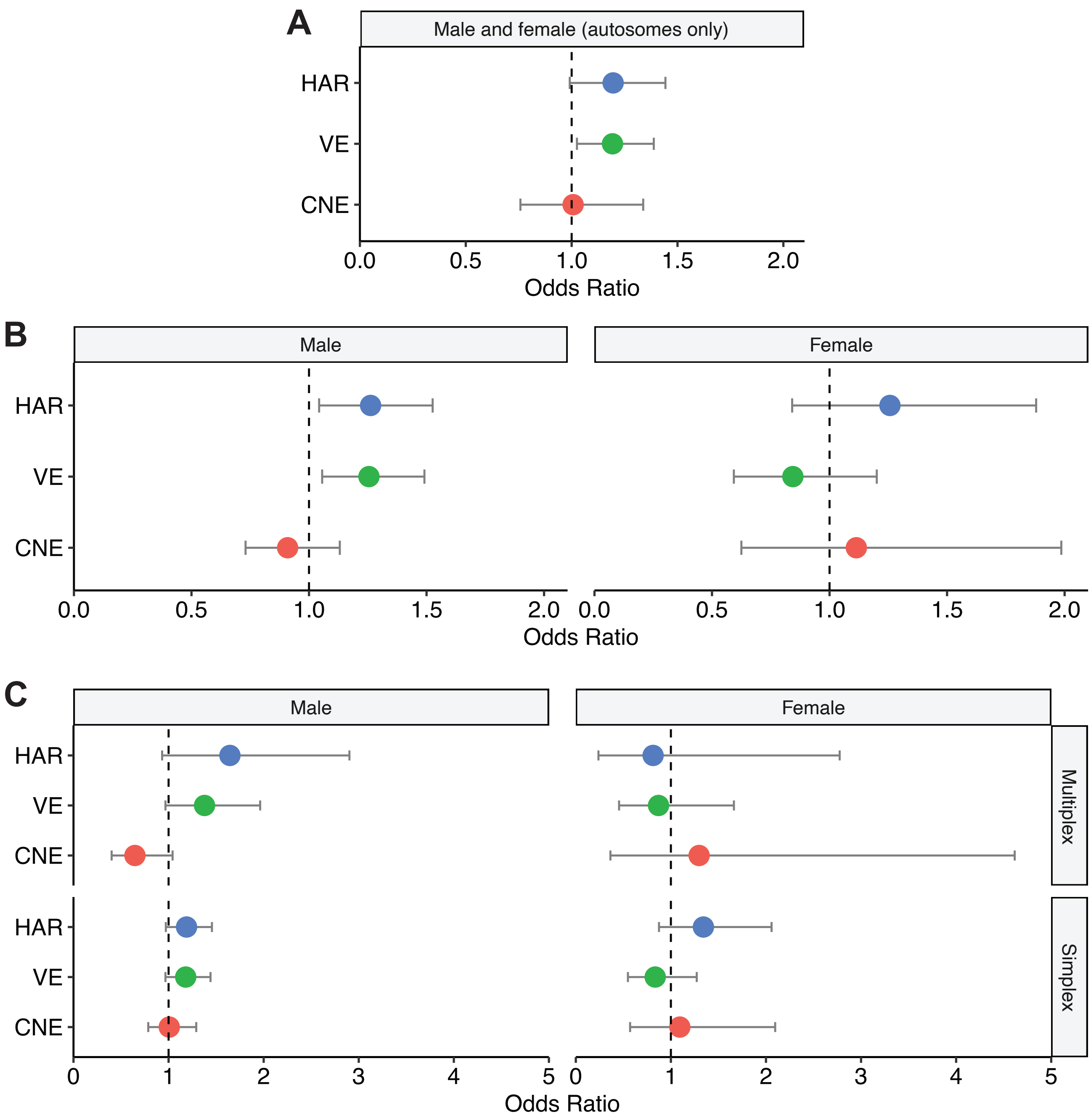


Figure S10: **Odds ratios for rare, recessive variants at conserved bases in cases versus controls in the NIMH cohort for (A) males and females (autosomes only), (B) males and females separately (autosomes and X chromosome), and (C) by family structure and sex (autosomes and X chromosome).** All are assessed at allele frequency (AF) < 0.001.

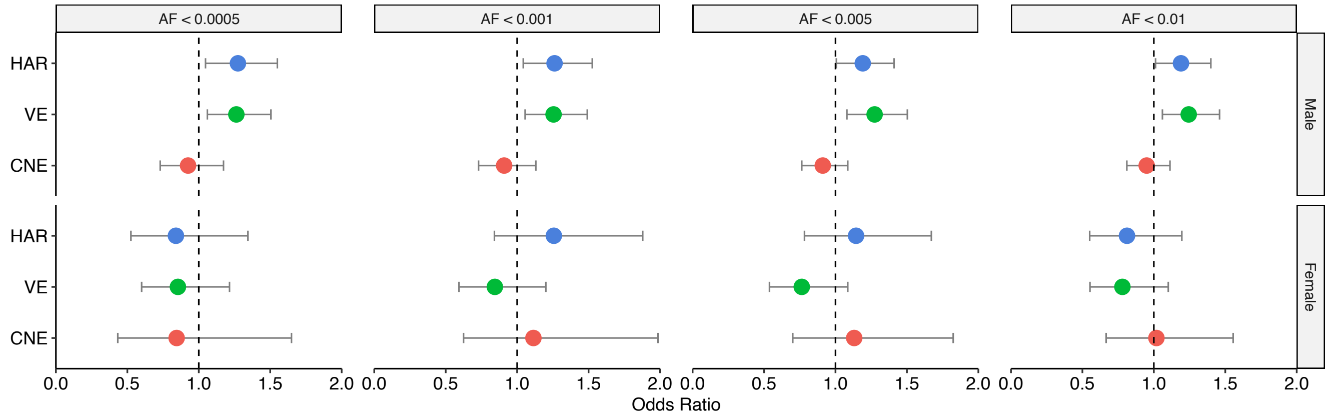


Figure S11: **Odds ratios for rare, recessive variants at conserved bases in cases versus controls in NIMH cohort are consistent across allele frequencies (AF).**

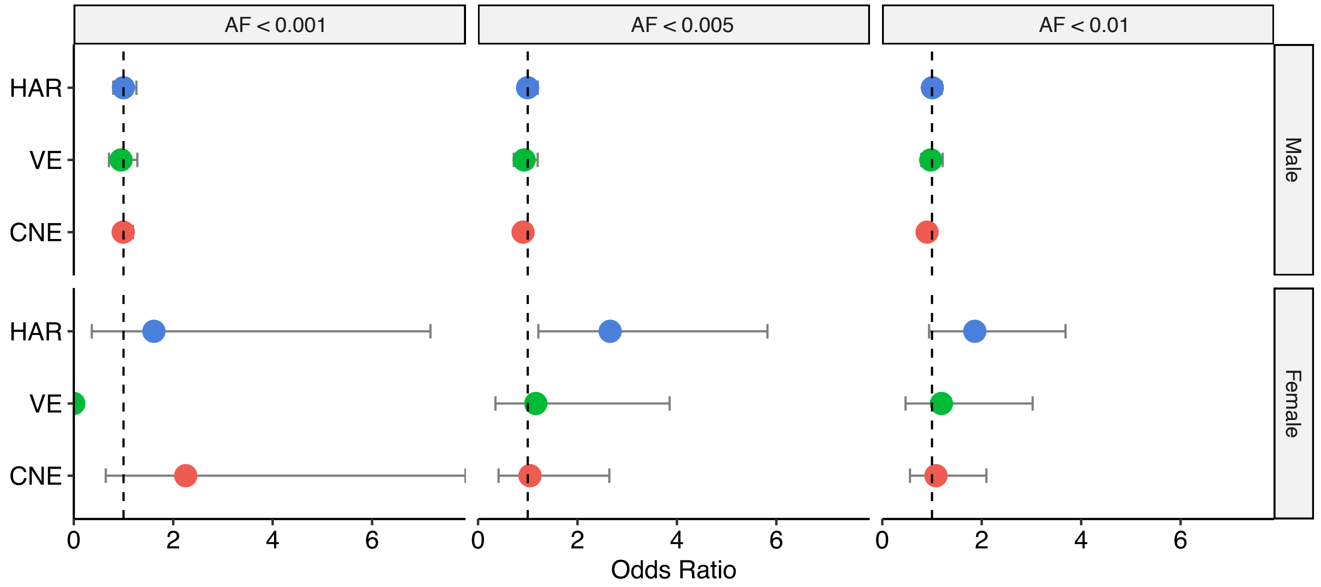


Figure S12: **Odds ratios for rare, recessive variants at conserved bases in cases versus controls in SSC cohort are consistent across allele frequencies (AF).**

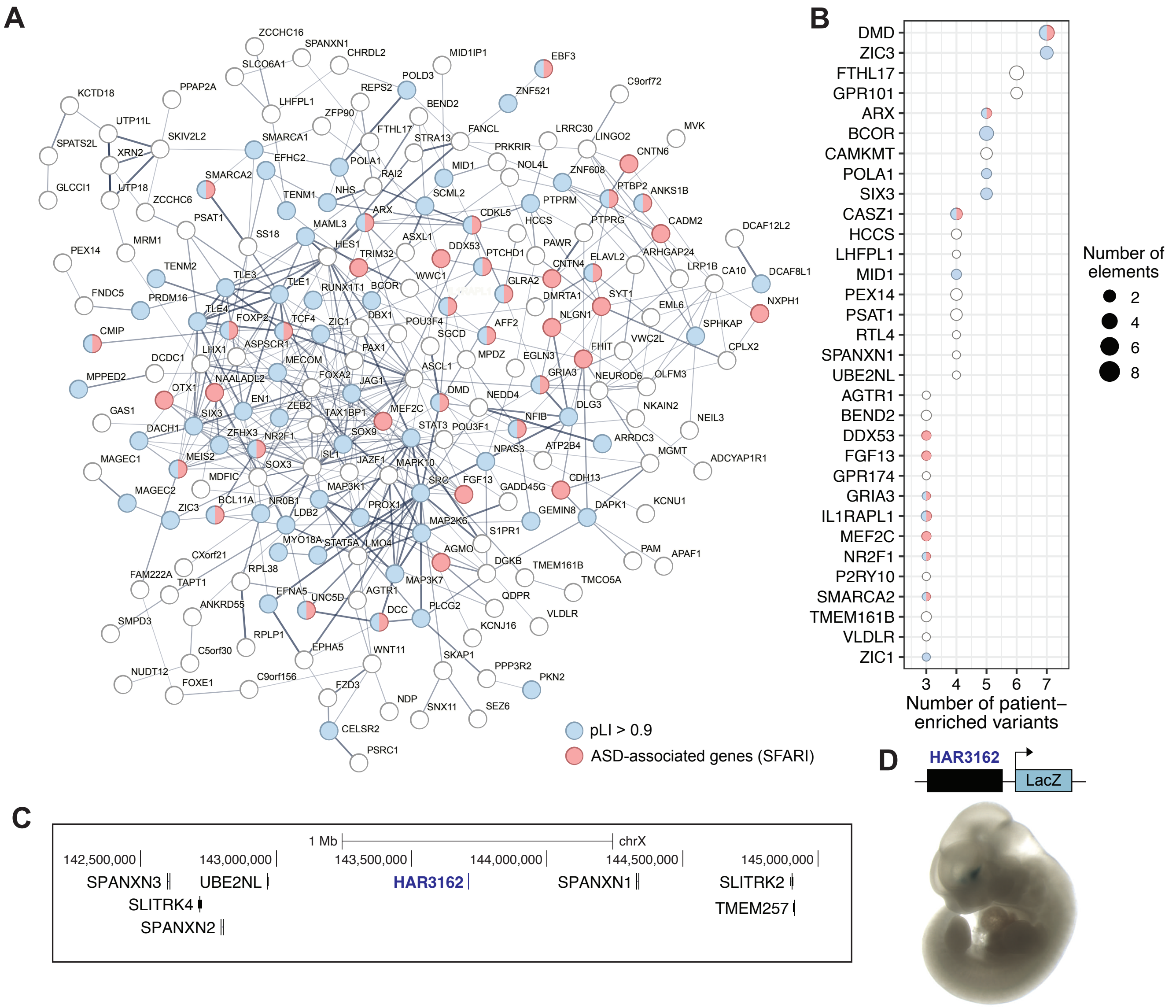


Figure S13: **Analysis of patient-enriched variants identified in the HMCA and NIMH cohorts.** (A) Protein-protein interactions of genes near HARs, VEs, and CNEs in the HMCA cohort and HARs and VEs in the NIMH cohort that have a numerical excess of variants found in cases compared to controls (Materials and Methods). Genes associated with ASD (Abrahams et al., 2013) are colored red, and genes that are loss-of-function intolerant (pLI > 0.9) (Lek et al., 2016) are colored blue. The thickness of network edges indicates the strength of data supporting the interaction, and only networks with >5 proteins were included. (B) The number of variants found in more cases than controls (patient-enriched variants) and the number of HARs, VEs, and CNEs that they are found in (elements) are plotted for genes near at least 3 patient-enriched variants. Only patient-enriched variants where the HAR, VE, or CNE they are located in has more patient-enriched than control-enriched variants are included. Genes associated with ASD (Abrahams et al., 2013) are colored red, and genes that are loss-of-function intolerant (pLI > 0.9) (Lek et al., 2016) are colored blue. (C) Genomic interval including HAR3162 and nearby genes. (D) HAR3162 cloned upstream of a minimal promoter driving the lacZ gene was integrated at the safe-harbor H11 locus and analyzed for lacZ expression at E11.5 (Materials and Methods). HAR3162 drives lacZ expression in the ventral telencephalon (representative embryo shown here).

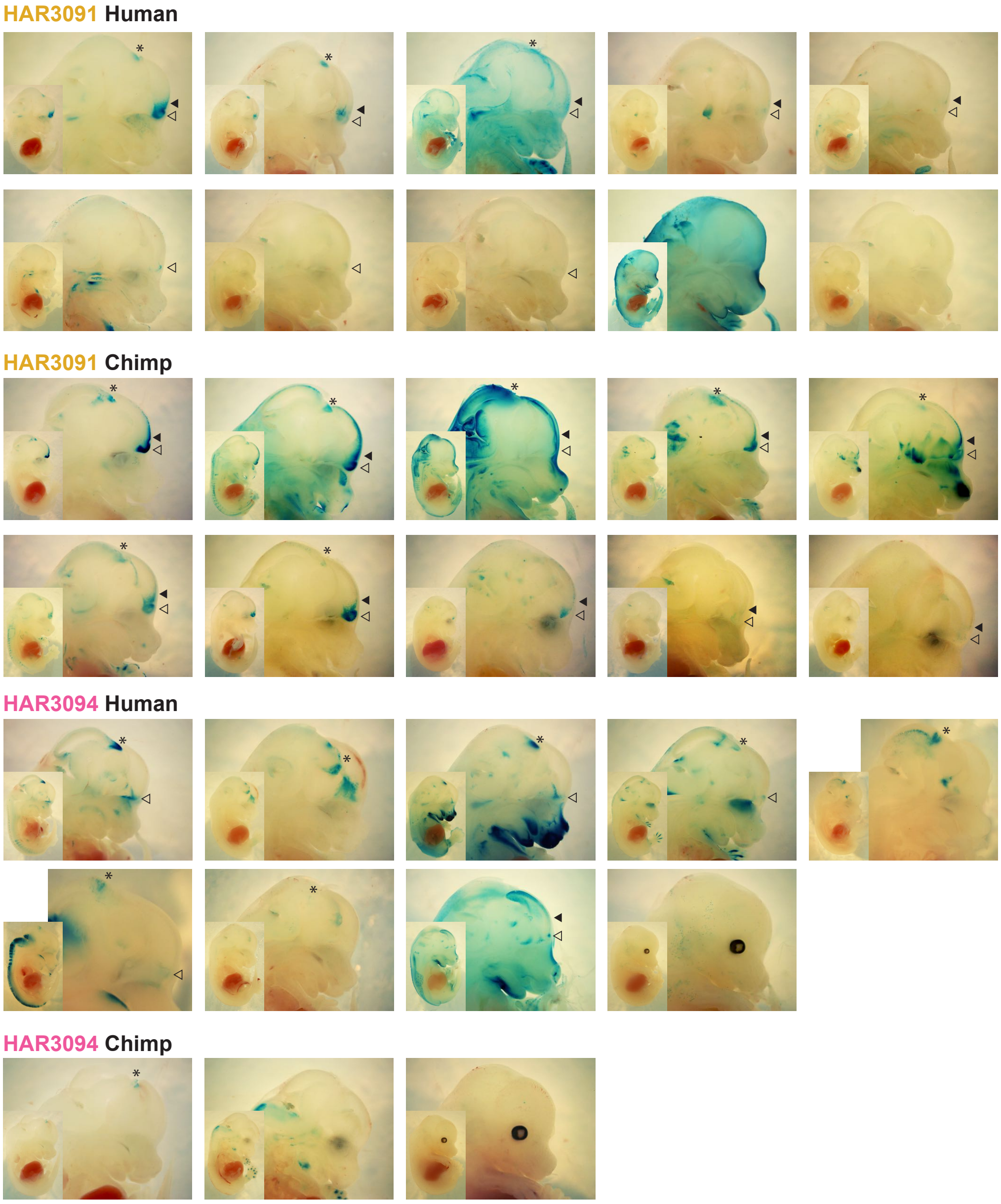


**Embryos with lacZ expression**

| | HAR3091 | | HAR3094 | |
|---|---|---|---|---|
| | Human | Chimp | Human | Chimp |
| telencephalon ◂ | 5/10 (50%) | 10/10 (100%) | 1/9 (11%) | 0/3 (0%) |
| olfactory bulb ◃ | 8/10 (80%) | 10/10 (100%) | 5/9 (56%) | 0/3 (0%) |
| midbrain * | 3/10 (30%) | 7/10 (70%) | 7/9 (78%) | 1/3 (33%) |

Figure S14: **Enhancer reporter assay of HAR3091 and HAR3094 in transgenic mice.** Enhancer reporter constructs containing the human or chimpanzee versions of HAR3091 and HAR3094 cloned upstream of a minimal promoter driving *lacZ* expression were injected into mouse embryos and analyzed at E14.5 (Materials and Methods). Embryos were genotyped for the *lacZ* gene from tail clips. There were 16 PCR-positive embryos for the human version of HAR3091, 14 PCR-positive embryos for the chimpanzee version of HAR3091, 15 PCR-positive embryos for the human version of HAR3094, and 10 PCR-positive embryos for the chimpanzee version of HAR3094. All embryos with any visible lacZ staining are displayed. Images are of bisected embryos, unless there was no internal lacZ staining. The table shows the percentage of embryos with lacZ expression that have expression in the telencephalon (filled arrowhead), olfactory bulb (unfilled arrowhead), or midbrain (asterisk). Visible lacZ staining is taken as a proxy that the full construct was integrated and is assessable in an embryo. Given the mosaic and random nature of integration events in this experiment, tissue regions where the tested sequences can drive enhancer activity will show staining in multiple embryos.

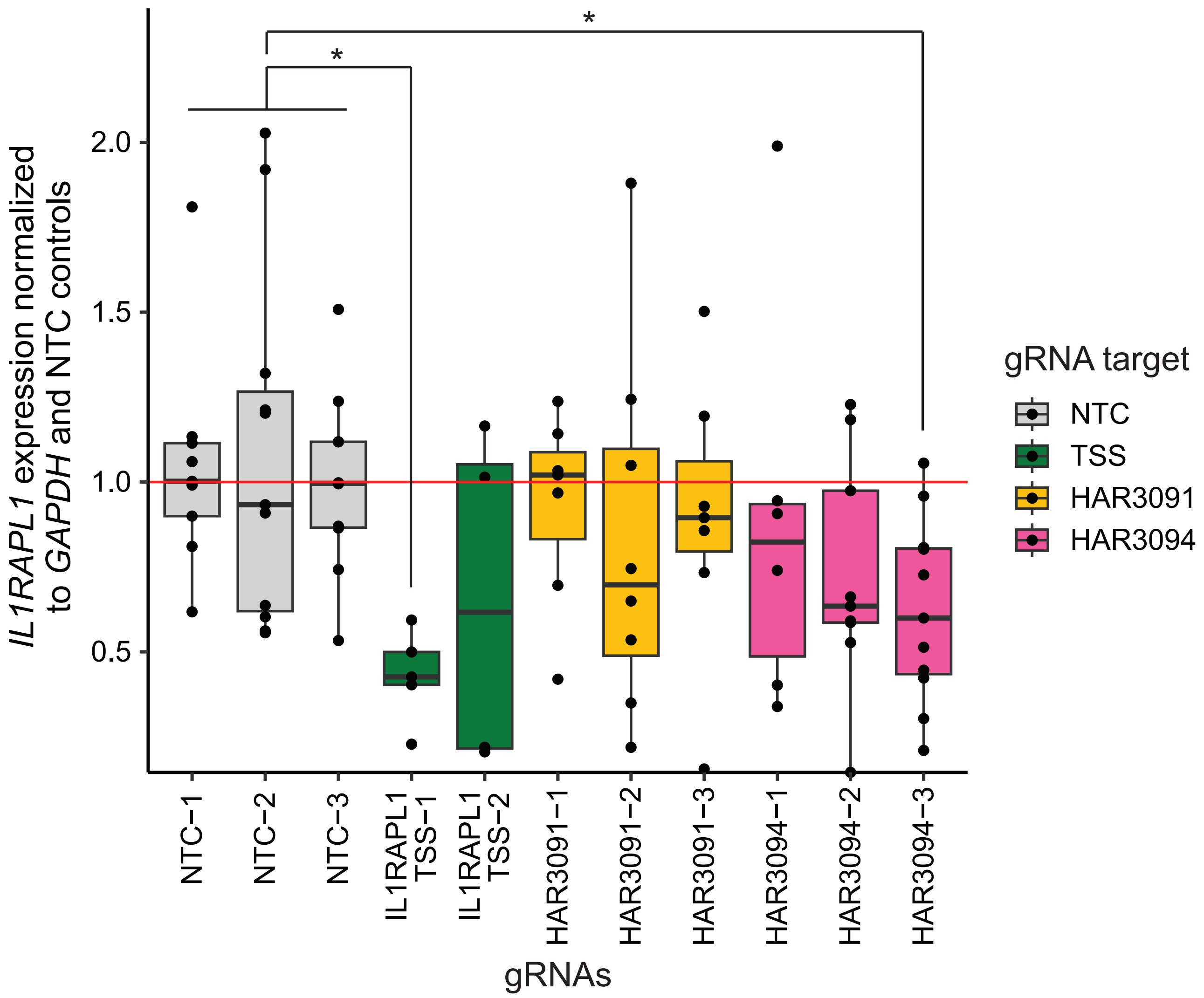


Figure S15: **CRISPRi targeting the *IL1RAPL1* promoter, HAR3091, and HAR3094.** We tested three non-targeting control (NTC) gRNAs (gray), two gRNAs targeting the *IL1RAPL1* promoter (green), 3 gRNAs targeting HAR3091 (yellow), and 3 gRNAs targeting HAR3094 (pink). Compared to the NTC gRNAs, only the gRNAs IL1RAPL1 TSS-1 (adjusted $p = 0.0002$) and HAR3094-7 (adjusted $p = 0.002$) were statistically significant.

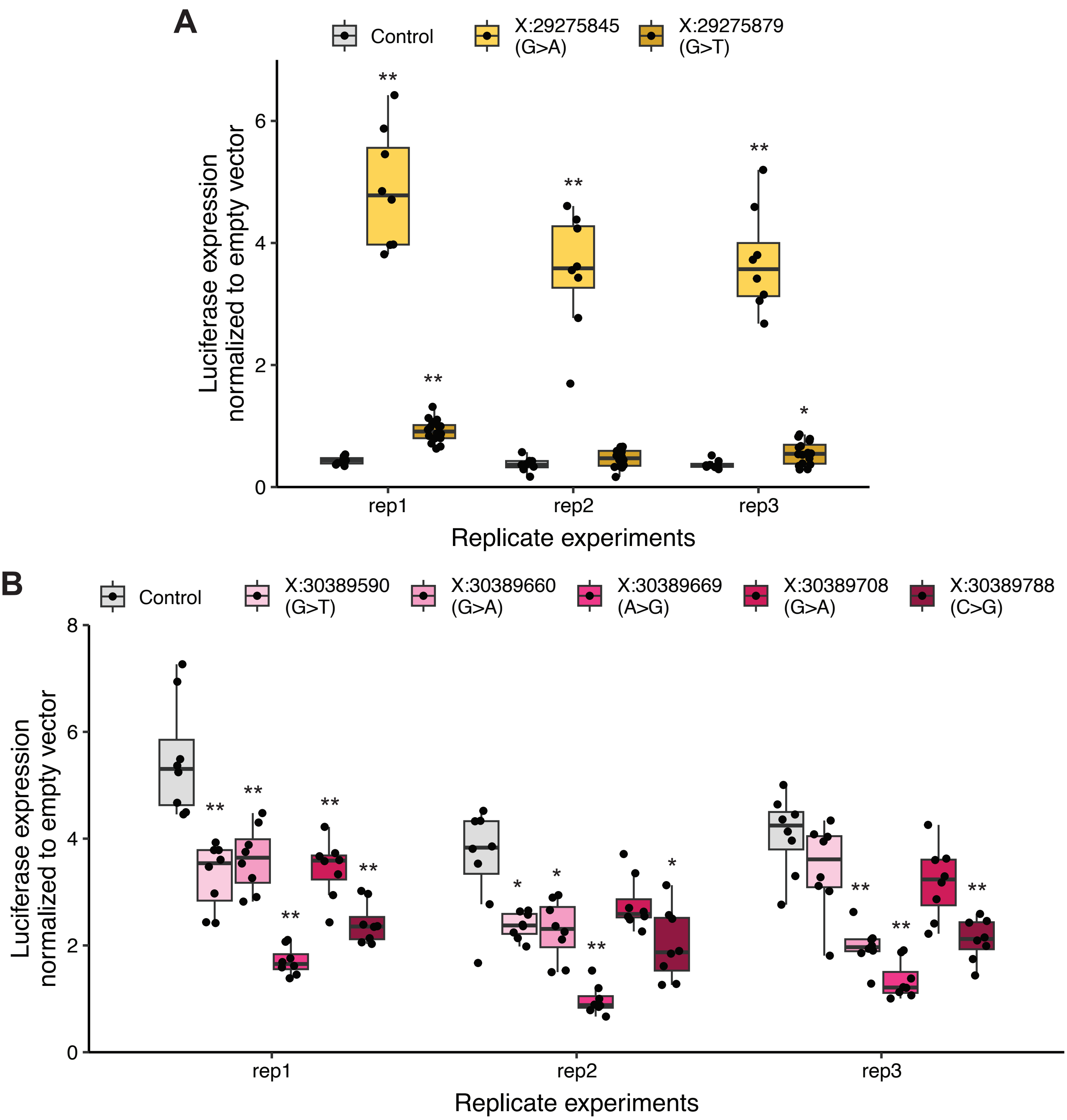


Figure S16: **Luciferase assays of patient variants in HAR3091 (A) and HAR3094 (B).** Within each replicate experiment, each patient sequence was compared to the luciferase expression from the control sequence with the Wilcoxon rank-sum test. P-values were adjusted using the Benjamini-Hochberg correction. * : adjusted $p < 0.1$, ** : adjusted $p < 0.01$.

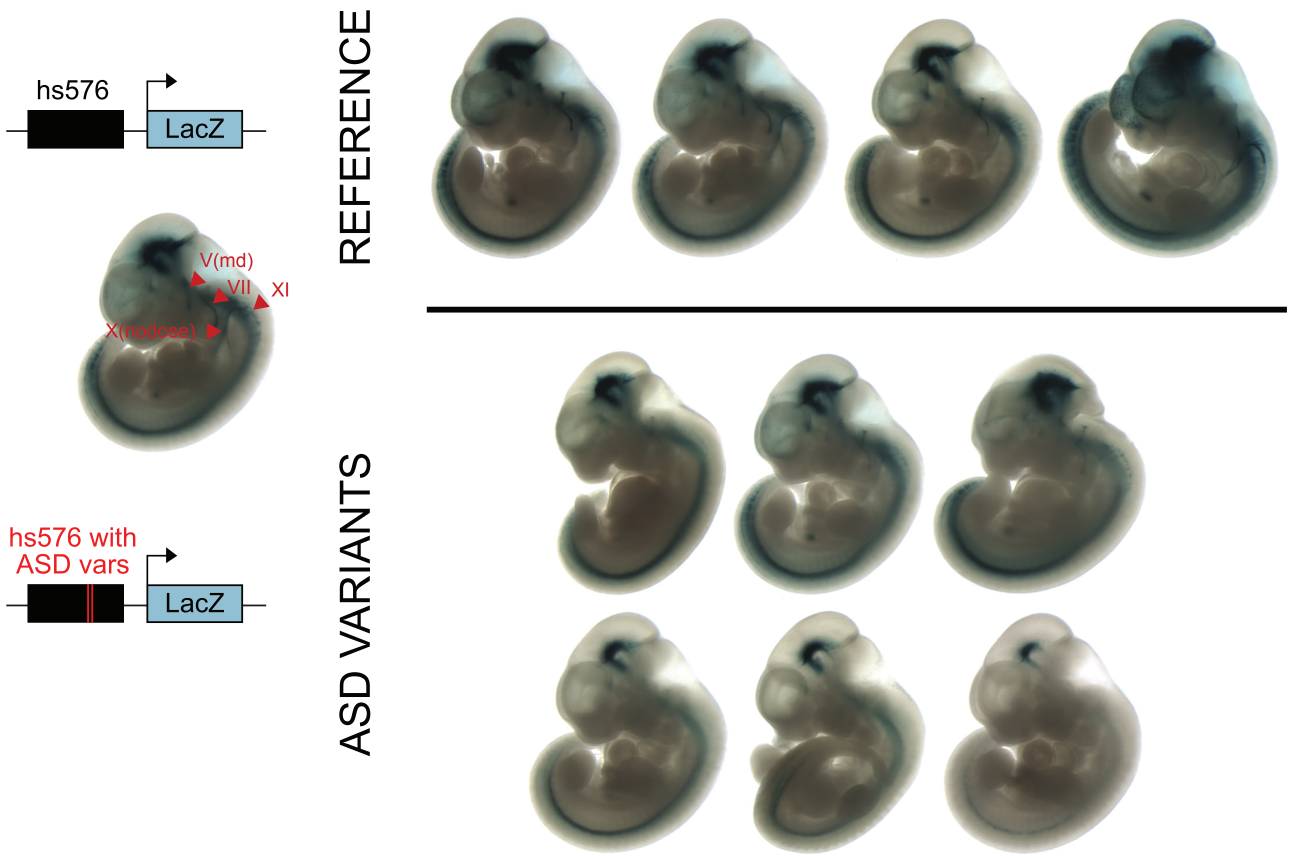


Figure S17: **Enhancer reporter assay of hs576 (VE854) in E11.5 transgenic mice.** Enhancer reporter constructs containing hs576 with or without the two ASD patient variants upstream of a minimal promoter driving lacZ expression were injected into mouse embryos, screened for stable integrants at the safe-harbor H11 locus, and analyzed at E11.5 (Materials and Methods). All embryos with homozygous insertions at the H11 locus are shown.

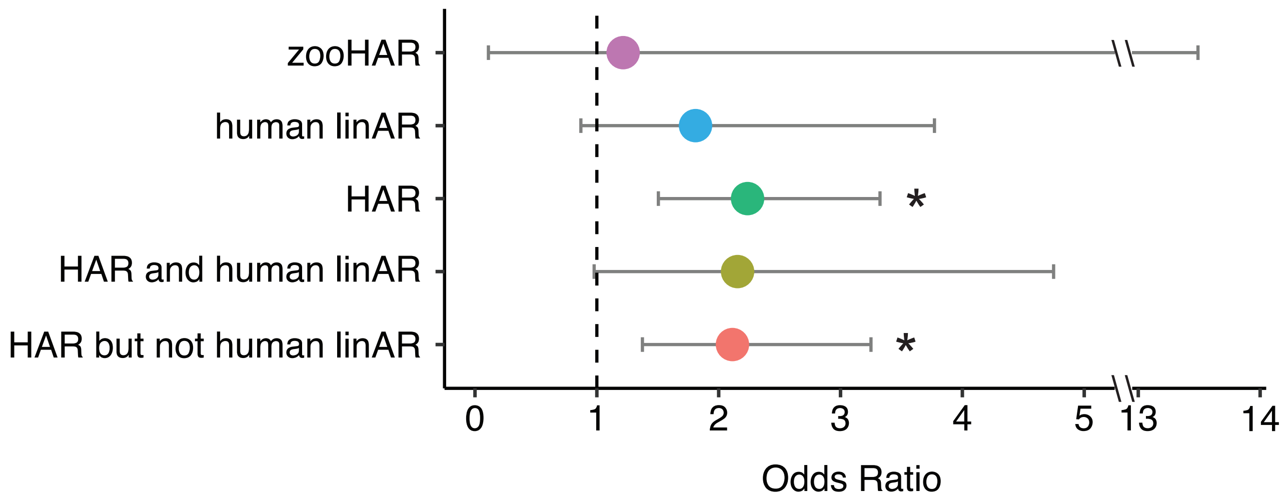


Figure S18: **Odds ratios for the number of rare, recessive variants at conserved bases in ASD cases compared to controls at allele frequency < 0.005 in the HMCA cohort for the set of HARs analyzed in this study (HAR) and two recently identified sets of HARs, zooHARs (Keough et al., 2023) and human linARs (Bi et al., 2023).** Given the moderate overlap between linARs and the HARs analyzed in this study, we also assessed HARs that overlap linARs (HAR and human linAR) and those that did not (HAR but not human linAR).

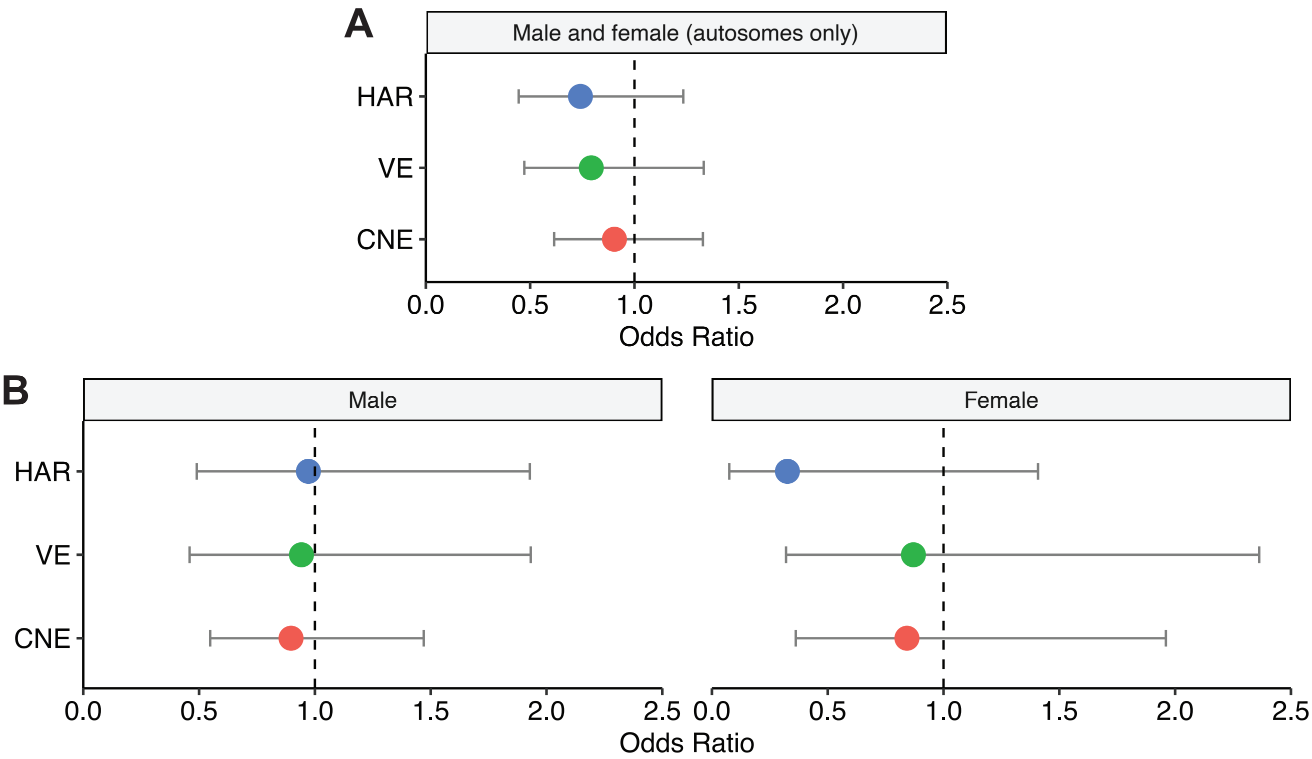


Figure S19: ***De novo* variants are not enriched in cases versus controls in SSC for (A) males and females (autosomes only) or (B) males and females separately (autosomes and X chromosome).**

### Supplemental Tables

Table S1: **Coordinates (hg19), TF motif enrichments, GREAT gene ontology results of HARs, VEs, and CNEs**
Table S2: **caMPRA results for HARs, VEs, and CNEs**
Table S3: **caMPRA results from random mutagenesis of HARs**
Table S4: **Rare, recessive variants at conserved bases in ASD cohorts**
Table S5: **Oligos used in this study**
